# The emergence of a novel synthetic nicotine analog 6-methyl nicotine (6-MN) in proclaimed tobacco- and nicotine-free pouches available in Europe

**DOI:** 10.1101/2024.10.31.24316471

**Authors:** Celine Vanhee, Maarten Dill, Michael Canfyn, Emmy Tuenter, Sophia Barhdadi

## Abstract

A new nicotine delivery system in the form of tobacco-free nicotine pouches, was introduced in Europe in 2019. These nicotine bags did not fall under the Tobacco Products Directive (TPD) II which brought forward regulatory requirements for both cigarettes, related products, and e-liquids. As these pouches did not fall under the scope of the TPD, it was up to the member states to decide which action to be taken if any. Some EU Member States banned these nicotine pouches, while others put restrictions on the amount of nicotine, warning messages, and presentation and packaging of the product, and some Member States did not take any action. Likely as a result of the ban or restrictions soon after also tobacco and nicotine-free pouches became available in the European Union (EU). Early 2024, “NoNIC” pouches, claiming to be tobacco- and nicotine-free, became available on the European market. These pouches are promoted online and clearly target a younger population with a youth-appealing package design and enticing flavors. Upon analysis of different samples utilizing gas chromatography coupled to MS (GC-MS), liquid chromatography coupled to high-resolution tandem mass spectrometry (HRAM LC-MS2) and nuclear magnetic resonance spectroscopy (NMR), it was demonstrated that these nicotine-free pouches contained the synthetic nicotine homolog 6-methyl nicotine (6-MN) and this up to 20 mg 6-MN per pouch for the highest strengths. Nonetheless, a part of a likely unaware/misled young adult subpopulation has easy access to these products, containing a novel molecule for which limited to no clinical effects are known.

## INTRODUCTION

Nicotine is a highly addictive molecule that acts, similarly to cocaine, alcohol, or opioids, on the brain by releasing the “reward” molecule dopamine and conveying mood-altering “feel good” changes [1–8]. For several centuries, the stems and the leaves of the plant *Nicotiana tabacum* or *Nicotiana rustica* were in use as the primary source of nicotine. Recent archeological findings have uncovered that the smoking of tobacco took place in North and South America more than 3000 years ago [9]. It was not until 400 years ago, that tobacco was introduced in Europe. Soon, the product became a global trade commodity and a mass consumable either in the form of a smoking product (e.g. cigarettes, pipe tobacco, cigars, cigarillo or hookah) or smokeless tobacco (e.g. chewing tobacco, sniffing tobacco or snus). At the beginning of the 20th century, reports started to emerge linking the prevalence of lung cancer and/or oral cavity cancer with the use of tobacco products [10,11]. It has been postulated that tobacco caused 100 million deaths in the 20th century [12]. Nonetheless, it took the European Union (EU) till 2001 to put in place the first EU Tobacco Products Directive (TPD), which regulated aspects such as the manufacture, sale, and presentation of tobacco products (directive 2001/37/EC) [13]. However, 8 years later, under the pressure of the public health community, and following scientific and market-related developments, the European Commission (EC) decided to review the TPD which was finalized in April 2014 [14, 15]. This TPD II also brought forward regulatory requirements for several design and packaging parameters for e-cigarettes, including maximum tolerated nicotine concentrations, child-resistant refill containers, and the regulation/notification of e-liquid constituents to regulators [16]. Recently, around 2019, a nicotine delivery system in the form of nicotine pouches was introduced in Europe, the USA, and Japan. These pouches resembled Swedish snus, a smokeless tobacco product, which is only allowed in Sweden but prohibited in any other EU country [14]. Nicotine pouches, also known as white snus, nicopods, and smokeless tobacco-free snus, consisting of a nicotine and a non-tobacco substance (e.g. microcrystalline cellulose or non-tobacco plant fibers) present in a teabag like sachet, called a pouch. Other ingredients, such as artificial sweeteners, additives, and flavorings, are also present and are sold in a variety of fruit and other flavors such as mint, muffin, and cola [17]. These pouches are promoted via several channels, including social media, and are often marketed as healthier than conventional cigarettes or e-cigarettes. These marketing strategies have also been used previously by e-cigarette manufacturers to entice young people. The usage of these pouches is not without risk as the product still contains nicotine, known to have detrimental effects on the still developing brain of youngsters and young adults and occasionally can also induce an intoxication in this partial age group, whether or not as a result of accidental ingestion [18–22]. Moreover, there has been data demonstrating that these pouches might potentially act as a stepping stone to cigarette smoking, the so-called gateway effect [23]. Based on these findings, some EU Member States, including Belgium, the Netherlands, Germany, and France banned the consumption and sales of nicotine pouches as they classified them as food products, where the acceptable daily intake (ADI) was set to 0.8 µg per kilogram bodyweight [24]. This corresponds to a maximum of 0.05 mg of nicotine a day for an individual with a 60 kg bodyweight. Alternatively, other Member States put restrictions on the products such as banning the sale to minors, limiting the amount of nicotine per pouch (e.g. 20 mg per pouch), and/or requiring the placement of warnings regarding the consumption of this product, while in other Member States these pouches are freely available [17]. Currently, a potential harmonization and even ban is under discussion during the current re-evaluation of the TPD II, resulting in TPD III, estimated to be finalized by the end of 2024 or early 2025.

However, as the legal restrictions evolve with scientific knowledge, so do some manufacturers evolve by searching for the Achilles’ heel in the regulatory framework. Indeed, in October 2023, the first report was made of the presence of an unregulated synthetic nicotine alkaloid analog, termed 6-methyl nicotine (6-MN), in an electronic cigarette pod system [25, 26]. At the time, 6-MN was not subjected to the 2022 newly amended U.S. tobacco and vaping regulation that regulated only nicotine-containing products independent of their origin (natural or synthetic) and did not cover methylated nicotine forms. This product was not marketed in the EU as it very likely would fall under the scope of the Tobacco Products Directive 2014/40/EU, applicable to electronic cigarettes, as nicotine is therein defined as nicotinic alkaloids [15]. However, in the beginning of 2024 tobacco-free and nicotine-free pouches, termed “NoNIC”, became available in Europe [27]. These pouches are easily available online through websites that employ different EU and UK-based domain names. Some of these sites claim only the absence of nicotine, while other shops state the presence of 6-MN. With this study, we set out to determine if these pouches were indeed devoid of nicotine and contained 6-MN, and if so, how much was present in them.

## 2. MATERIALS AND METHODS

### 2.1. Solvents and chemical reference standards

Mass spectrometry (MS)-grade acetonitrile (purity 99.8%), methanol (purity > 99.9%), water, and formic acid (purity > 99%) were purchased from Thermo Fisher Scientific (Waltham, MA, USA). Deuterated methanol (>99.8% D) was obtained from Sigma-Aldrich (Steinheim, Germany). Reference material for nicotine (purity > 99%) and 6-methyl nicotine (purity = 92%) was purchased from Merck (Darmstadt, Germany) and Toronto Research Chemicals (North York, Ontario, Canada), respectively. Stock solutions were made in methanol at a concentration of 1 mg/mL and then further diluted into working concentrations either for confirmation of the screening result (dilution with methanol) or for quantification purposes (dilution with water).

### 2.2. Samples

A total of nine samples, covering the three different strengths and eight different flavors were analyzed. Three of the nine samples were collected by the Federal Public Service Health, Food Chain Safety, and Environment and sent to our laboratory to screen for nicotinic alkaloids. These samples represented the flavor, mint, and the three available strengths: super kick or extra strong, double kick or super strong. Six additional samples, representing different strengths and flavors, were purchased online in July 2024 at two different UK-based vendors. Upon arrival, the samples were kept closed at room temperature 15-25°C and were opened for a maximum of one week until quantification.

### 2.3 Screening for nicotine alkaloids or other ingredients by MS

Prior to the MS analysis, the content of 3 pouches was mixed and approximately 25 mg of this mixture was weighted and resuspended in 10 mL methanol, followed by a sonication for 10 minutes and a filtration through a 0.2 µm PTFE filter. The collected filtrate was then subjected to gas chromatography coupled to MS (GC-MS) and liquid chromatography coupled to high-resolution tandem mass spectrometry (HRAM LC-MS2).

The GC-MS analysis was performed on an Agilent 7890A gas chromatograph coupled to an Agilent 5975C mass detector (Agilent, Agilent, Santa Clara, CA, USA) as previously described [28]. Briefly, all injections were performed in pulsed spitless mode, with an injection port temperature of 250 °C. The solvents were separated in an Agilent DB-5MS Agilent DB-5MS (40 m x 0.25 mm x 0.25 µm film thickness). The column oven temperature was initially set to 80 °C for 2 min and then raised at a rate of 15 °C/min to 280 °C and held for 17 min, followed by an increase of 10 °C/min to 310 °C and held for 20 min. The total run time was 55 min. High-purity helium was used as the carrier gas with a flow rate of 1 mL/min. The MS was operated in electron ionization mode with an electron energy of 70 eV. Data were acquired in full-scan mode with m/z ranging from 43 to 560. The obtained GC-MS data were analyzed by MassHunter and the spectra were compared to different libraries, including the National Institute of Standards and Technology (NIST 20) mass spectral library (NIST, Gaithersburg, MD, USA) and the Cayman spectral library version 30052024 (Cayman Chemical, Ann Arbor, Michigan, USA).

The samples were also simultaneously analyzed by high-resolution accurate mass (HRAM) tandem MS (MS/MS). The analyses were carried out on a Thermo Scientific™ Vanquish™ ultra-high performance liquid chromatography (UHPLC) system coupled to a Q-Exactive™ focus orbitrap mass spectrometer equipped with a heated electrospray ionization (ESI) source operating in positive ion mode (Thermo Fisher Scientific, Bremen, Germany). The LC method used for nicotine alkaloids, employing an Acquity UPLC HSS T3 column (100 mm × 2.1 mm i.d., 1.8 μm particle size) (Waters, Milford, MA, USA), was as follows: 0–1 min 100% A (0.1% formic acid in water), 6 min 30% B (0.1% formic acid in acetonitrile), increased to 99% B in the next 0.1 min, then was kept at 99% B for 1.4 min, followed by a 1.5 min re-equilibration step (total run time: 9 min). The flow rate was set at 0.4 mL/min, the column temperature at 40 °C, and a 2 µL sample or reference standard was injected. The high-resolution accurate mass analysis was performed on a Q-Exactive Focus Orbitrap MS, equipped with a heated electrospray ionization (ESI) source operating in positive ion mode. The optimized Orbitrap tune method was set as follows: the flow rate of the sheath gas (nitrogen, purity ≥ 99.99%) and the auxiliary gas (nitrogen, purity ≥ 99.99%) was set at 30 and 10 arbitrary units, respectively; the temperature of the capillary was set at 320 °C; the voltage of the spray was 3.2 kV; and the S-lens RF level was set at 60 V. The MS scan data were acquired at a resolving power of 35,000 (at m/z 200) while the MS2 data were recorded with a resolving power of 17,500. The collision energy for the data-depended MS2 (ddMS2) of the precursor ion was adjusted to 30 CE. All data were acquired by using the Thermo Xcalibur 4.0 software and processed using FreeStyle™ 1.7 SP2 software (Thermo Fisher Scientific, Bremen, Germany).

A possible match with a nicotine alkaloid or 6-MN was verified by means of an injection with a reference standard at a concentration of 0.05 mg/mL. For the GC-MS data, a compound was considered present when a) the retention time shift was less than or equal to 0.2 min (compared to the reference standard), b) there was an 85% or greater match of the fragment ions and their relative intensities between the MS spectrum of the sample and the MS spectrum of the reference standard. For the LC-MS2 analysis, a compound was considered present provided that a) the difference in retention was also less than or equal to 0.2 min (compared with the retention time of the reference standard of this compound), b) the m/z of the precursor ion was equal to the one obtained with the reference standard (error tolerance of 10 ppm), c) and the MS2 spectra of the observed fragment ions concurred with those obtained for the reference standard (precursor and fragment ions and their corresponding relative intensities are given in Table 1). The acceptable relative errors on the relative intensities of the fragment ions were as follows: for relative intensities between 10 and 40:30%; between 40 and 60:25%; and above 60 a relative error of 10% was deemed acceptable.

**Table 1.** The chemical formula, the comparison of the theoretical and observed mono-isotopic mass of the precursor and main fragment ions of both reference standards that were analyzed by HRAM LC-MS, and their relative intensities.

| The chemical formula of precursor and fragment ions | Calculated monoisotopic mass | Observed mono- isotopic mass | Mass difference ($\Delta$ ppm) | Intensity in MS2 (%) |
| --- | --- | --- | --- | --- |
| **R=H: nicotine (C<sub>10</sub>H<sub>14</sub>N<sub>2</sub>)** |  |  |  |  |
| C<sub>10</sub>H<sub>15</sub>N<sub>2</sub><sup>+</sup> | 163.1220 | 163.1217 | -1.84 | 100 |
| C<sub>9</sub>H<sub>10</sub>N<sup>+</sup> | 132.0808 | 132.0799 | -6.81 | 60 |
| C<sub>9</sub>H<sub>8</sub>N<sup>+</sup> | 130.0651 | 130.0644 | -5.38 | 20 |
| C<sub>7</sub>H<sub>8</sub>N<sup>+</sup> | 106.0651 | 106.0647 | -3.77 | 25 |
| C<sub>5</sub>H<sub>10</sub>N<sup>+</sup> | 84.0808 | 84.0806 | 7.14 | 20 |
| C<sub>5</sub>H<sub>6</sub>N<sup>+</sup> | 80.0495 | 80.050 | 6.25 | 10 |
| **R=CH3: 6-methyl nicotine (C<sub>11</sub>H<sub>16</sub>N<sub>2</sub>)** |  |  |  |  |
| C<sub>11</sub>H<sub>17</sub>N<sub>2</sub><sup>+</sup> | 177.1386 | 177.1376 | -5.65 | 100 |
| C<sub>10</sub>H<sub>12</sub>N<sup>+</sup> | 146.0964 | 146.0957 | -4.79 | 60 |
| C<sub>10</sub>H<sub>10</sub>N<sup>+</sup> | 144.0808 | 144.0802 | -4.16 | 20 |
| C<sub>8</sub>H<sub>10</sub>N<sup>+</sup> | 120.0808 | 120.0802 | -5.00 | 35 |
| C<sub>6</sub>H<sub>8</sub>N<sup>+</sup> | 94.0651 | 94.0650 | -1.06 | 20 |
| C<sub>5</sub>H<sub>10</sub>N<sup>+</sup> | 84.0808 | 84.0808 | / | 20 |

### 2.4 Confirmation of the presence of 6-MN by NMR spectroscopy

A methanol extraction was performed on two pouches from the strength tiple kick and the flavor “mango” as fewer peaks were visible in GC-MS for this flavor compared to the other triple kick flavors. These pouches were extracted with 50 mL of dry methanol by shaking (180 rpm, 30 min) followed by a centrifugation step at 1000 x g for 5 min. The extract was then subsequently concentrated and dried through a SpeedVac vacuum concentrator (Eppendorf, Hamburg, Germany), prior to cold shipment (-20°C) for NMR analysis. The sample, as well as athe reference compound, were dissolved in 600 μL methanol-*d*_4_ and submitted to NMR analysis on an Avance Nanobay III NMR spectrometer (Bruker, Rheinstetten, Germany), operating at 400 MHz for ^1^H-NMR and at 100 MHz for ^13^C-NMR experiments. ^1^H, ^13^C, as well as 2D (COSY, HSQC, HMBC) NMR spectra, were recorded.

### 2.5 Quantification of 6-MN

The amount of 6-MN was determined on a Waters Acquity UPLC™ system (Waters Corp., Milford, MA, USA) including a binary solvent manager, sample manager-flow through needle, column heater, and photo-diode array (PDA) detector connected to a Waters Empower 3.7.0 data station. The chromatographic separation was performed as described by Barhdadi, who developed and validated the method for the quantification of nicotine and nicotinic alkaloids in e-liquids [29]. Briefly, an Acquity™ UPLC BEH C18 Column (100 × 2.1 mm, 1.7 μm particle size) (Waters, Milford, MA, USA) was used in combination with a mobile phase consisting of 10 mM ammonium borate pH 9.0 (A) and acetonitrile (B). The developed elution method was employed as follows: 5% B held for 1 minute, followed by a linear gradient to 25% B over 6 minutes and kept for 2 minutes, followed by a return to 5% held for 2 minutes for re-equilibration (total run time was 11 minutes). The flow rate was set at 0.4 ml/ min, the column temperature was 30 °C and the samples and reference standards were kept at 10 °C. 6-MN was monitored at a wavelength of 260 nm. The samples were prepared as described in Digard et al.,2013 [30]. Whole pouches were extracted with 50 mL of dry methanol by shaking (180 rpm, 30 min) followed by a centrifugation step at 1000 x g for 5 min. A small aliquot of the sample was then further diluted in 10 mM ammonium borate pH 9.0 to attain a theoretical concentration (based on the indirect indication of the amount of 6-MN per pouch) of approximately 10 µg/mL as a mini-validation was used to demonstrate the suitability of the method for concentrations from 5-20 µg/mL. A method accuracy of 5% was deemed fit for purpose. Peak identity for 6-MN was assigned by analysis of a reference standard, comparing the retention time and ultra-violet absorbance spectra. The purity angle and purity threshold were checked for all samples and were found to be below the purity threshold, indicating a good purity of the chromatographic peak.

The quantification of the samples was performed on 3 separate pouches. All preparations (n=3) were injected 4 times and these 12 injections were used to generate the mean amount of 6-MN present in the sample. The measurement uncertainty was estimated as a confidence interval using the standard deviations obtained for these preparations.

## RESULTS

### 3.1 Ease of access to products and visual inspection of the packaging

Upon the entry of “NoNIC pouches” into the Google search engine, several hits were found for online shops with EU and UK-based domain names that delivered to Belgium. These pouches were available in three different strengths, super kick-extra strong, double kick-super strong, and triple kick-mega strong, and with different flavors (muffin, cola ice, candy tobacco, double mint, freeze ice, menthol ice, mango ice, peach ice, exotic ice, blueberry ice, ruby berry ice, and rainbow drops). In early July 2024, we purchased different strengths and flavors from three different web shops, and two out of the three were able to provide us with the requested product, demonstrating the ease of purchasing these items. The items passed customs with much ease as the packages were declared to contain candy or earplugs. In total, 6 different samples were purchased and supplemented with 3 different samples that were collected by the Federal Public Service Health, Food Chain Safety and Environment. Prior to the chemical analysis of these samples, pictures were taken of the front and the back of the different samples and their content. As can be seen in Figure 1, illustrating the different strengths and flavors, the front of the boxes states the presence of 25 “NoNIC” pouches, while the label on the back of the boxes clearly states the absence of nicotine but labels the GHS06 symbol and “danger” and for some samples the phone number of the Belgian anti-poison center was also present. Icons demonstrating age restrictions (18+) or pregnancy-related warnings were depicted on the side of the boxes. Interestingly, the label of all these samples mentioned a compound named “(S)2-methyl-5-(1-methyl-2-pyrolidinylo)pyridine”, in addition to the flavoring substances. This compound closely resembles “2-methyl-5-[(2S)-1-methylpyrrolidin-2-yl]pyridine”, the IUPAC name for 6-MN. No information could be found on the box about the amount of this compound present in the samples, nor was such information available on the brand website or the vendor’s website (see supplemental Figure 1A). Either the amount of nicotine was expressed or the amount of “NoNic” present in the sample was mentioned. Moreover, it was sometimes, unclear if the amount of 6-MN was expressed per gram of product or per pouch (see supplemental Figures 1B and C).

**Figure 1.**
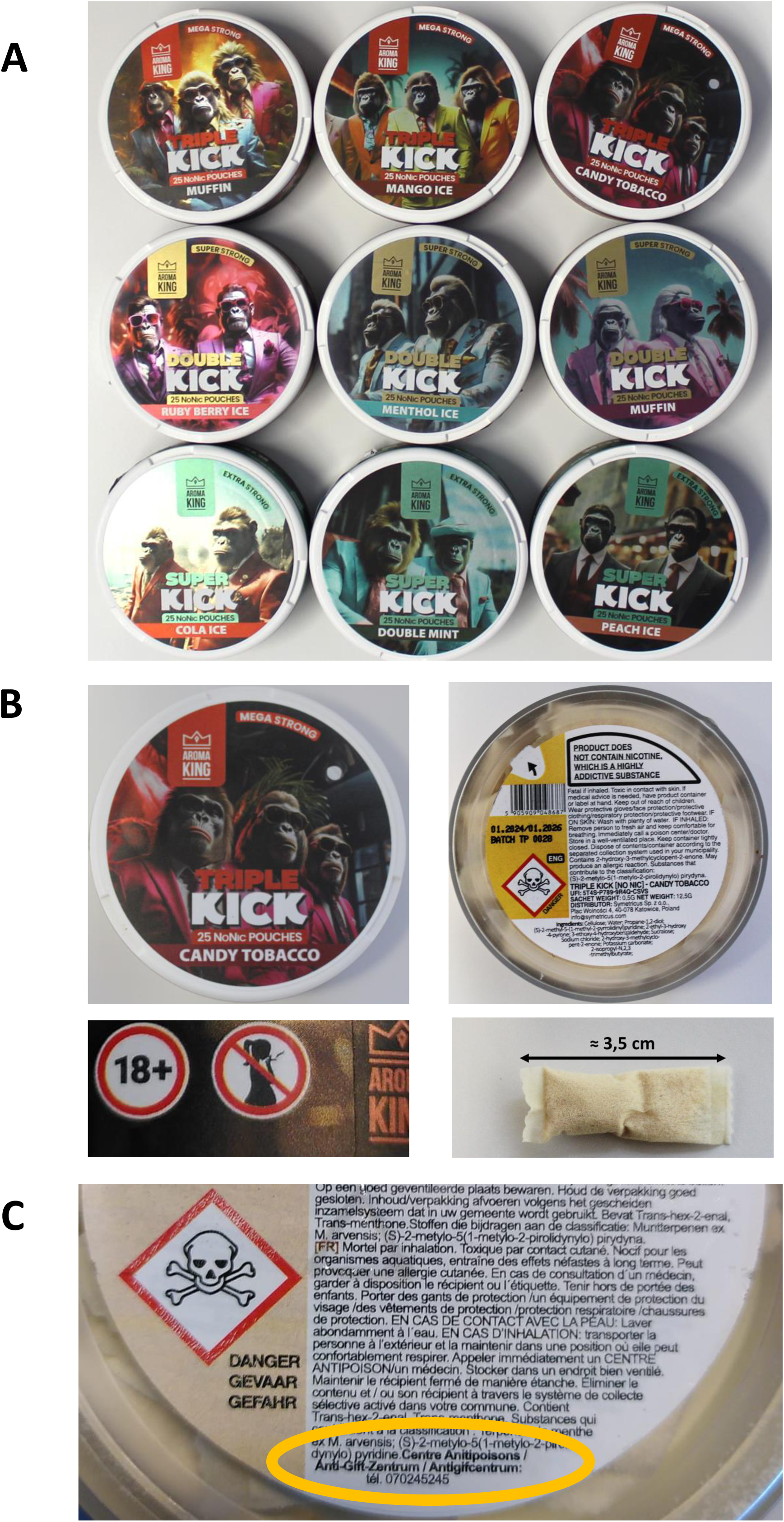
Pictures were taken from the boxes of the 9 different samples, representing different strengths and flavors (A). A close-up was also made of the front and back of one specific sample, the warning pictograms present on the side, and a pouch. The samples that originated from the Federal Public Service Health, Food Chain Safety and Environment also displayed the phone number of the Belgian anti-poison center.

### 3.2 Screening for nicotine and identification of 6-MN

Upon analysis of the content of the pouches by GC-MS and HRAM LC-MS^n^, also the reference standards of both nicotine and 6-MN were analyzed by the same method. The chemical structures, chemical formula, and the respective CAS number of the two reference standards are depicted in Figure 2A. The GC-MS spectra generated for the reference standards and the samples can be seen in Figure 2B. Nicotine eluted at 7.9 minutes during the GC-MS analysis, while 6-MN and the compound present in the samples eluted at 8.5 minutes. Moreover, the spectrum of nicotine showed the occurrence of the fragment ion 162.2 with 35% relative intensity, while this ion was absent in the spectrum of 6-MN and in the spectrum the compound, present in the samples. In fact, the spectrum obtained from this compound was almost a perfect overlay of the spectrum obtained for 6-MN. A similar phenomenon was also observed for the MS2 data generated by the HRAM LC-MSn analysis of 6-MN, as shown in Figure 3A. All compounds eluted at 0.7 minutes, but differences could be observed in the MS and MS2 spectrum of nicotine and 6-MN, the latter having an almost identical MS2 spectrum as the compound present in the pouches. The spectrum of nicotine showed the presence of the typical fragment ions visible in Figure 3A. A comparison of the theoretical and observed m/z values of these fragment ions of nicotine and 6-MN can be found in Table 1. Most of these fragment ions of 6-MN and the compound present in the pouches differed by an m/z value of 14, corresponding to a methylation of the pyridine group as can be seen in Figure 2C and Figure 3C. However, as several possible methylation sites are possible, NMR analysis was performed on an extract of a triple kick pouch in order to determine the methylation position. In the downfield region of the 1H-NMR spectrum of the pouch extract, three signals, each corresponding to one aromatic proton, were observed: one doublet (2.2 Hz) at 8.36 ppm; one double doublet (8.1; 2.2 Hz) at 7.75 ppm and one doublet (8.1 Hz) at 7.31 ppm. These multiplicities and J-values are typical for an aromatic ring with an ABX pattern, and thus are consistent with 6-MN, but not with 5-MN. Further confirmation was provided by a comparison of the ^1^H-NMR spectrum of the pouch extract with the reference compound. Although the extract is a mixture, all signals present in the spectrum of the reference compound could be observed in the pouch extract spectrum as well. Previously, the 1H-NMR data of 6-MN in CDCl_3_ or in CCl_4_ were reported [31, 32], but NMR data in methanol-d_4_ and/or ^13^C-NMR assignments were not found in the literature. Thus, additionally, ^13^C and 2D-NMR spectra were recorded as final confirmation to allow for the complete assignment of ^1^H- and ^13^C-NMR data. Again, the data obtained for the pouch extract were an exact match with those obtained for the reference compound. The NMR data can be found in Table 2. The NMR spectra can be consulted in the supplementary data Figures S2-S19.

**Figure 2.**
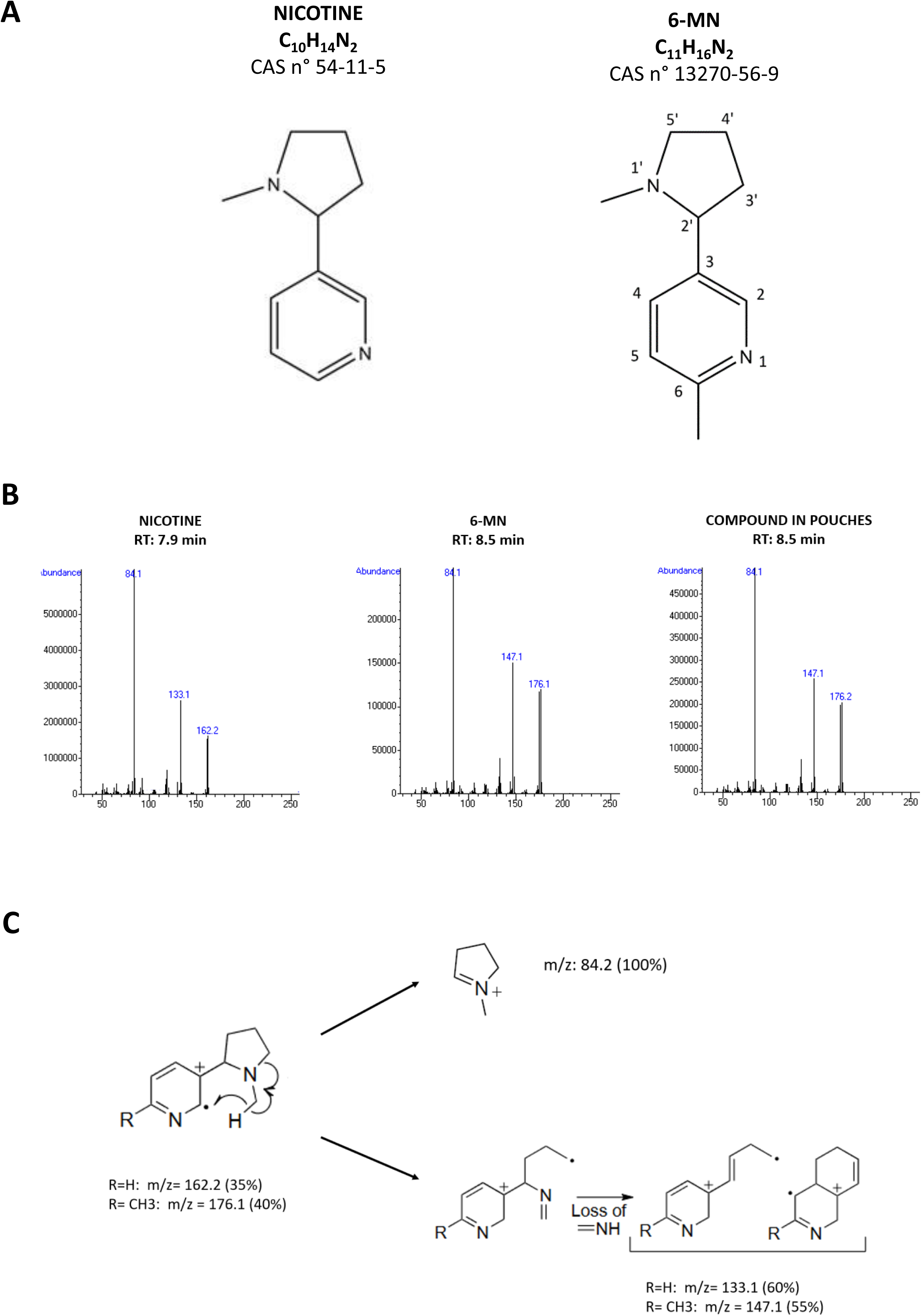
The structure of nicotine and 6-MN (A), the obtained GC-MS spectra of both reference standards and the compound present in the pouches (B), and the main fragmentation reactions (C). Nicotine eluted with a retention time of 7.9 minutes while 6-MN and the compound present in all samples eluted at 8.5 minutes.

**Figure 3.**
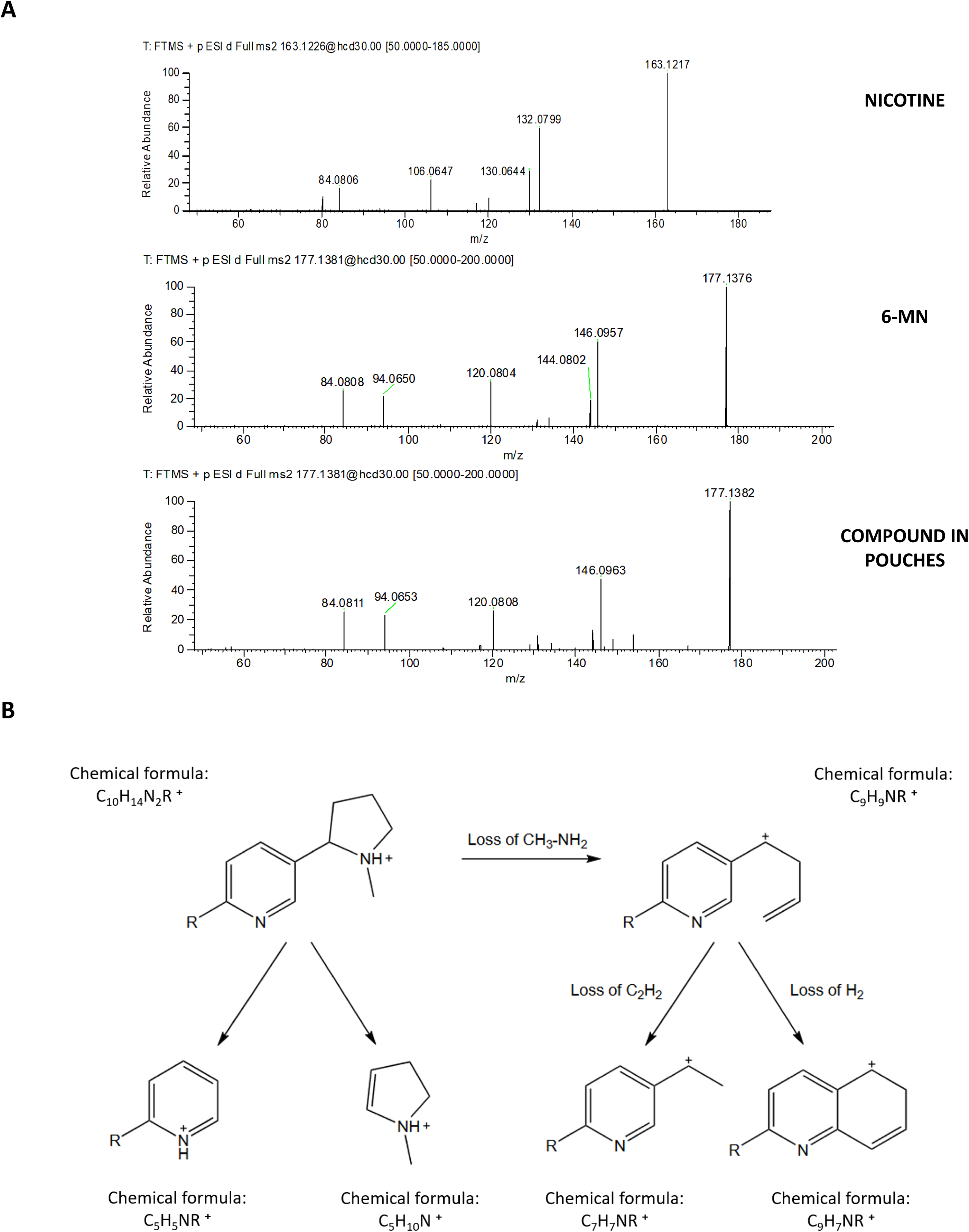
The LC-MS2 spectra of nicotine, 6-MN, and the compound present in the pouches (A) and the main fragmentation reactions (B).

**Table 2.** ^1^H- and ^13^C-NMR spectroscopic data of 6-MN (solvent: methanol-*d*_4_); δ in ppm, multiplicity (J in Hz). Spectra were recorded at 400 MHz for 1H and 100 MHz for 13C.

|  | <sup>1</sup>H |  | <sup>13</sup>C |
| --- | --- | --- | --- |
| position | $\delta_{\text{H}}$ in ppm,<br>multiplicity (J in Hz) | integration | $\delta_{\text{C}}$ in<br>ppm, type |
| 2 | 8.36, d (2.2) | 1 | 148.9, CH |
| 3 | - |  | 136.4 |
| 4 | 7.75, dd (8.1; 2.2) | 1 | 137.6, CH |
| 5 | 7.31, d (8.1) | 1 | 124.9, CH |
| 6 | - |  | 158.3 |
| 6-Me | 2.53, s | 3 | 23.2, CH<sub>3</sub> |
| 2' | 3.19, dd (9.2; 7.9) | 1 | 69.8, CH |
| 3' | 2.25, m ; 1.80, m | 1 / 1 | 35.2, CH<sub>2</sub> |
| 4' | 1.99, m; 1.90, m | 1 / 1 | 22.9, CH<sub>2</sub> |
| 5' | 3.25, m ; 2.38, q (9.2) | 1 / 1 | 57.7, CH<sub>2</sub> |
| N-Me | 2.18, s | 3 | 40.3, CH<sub>3</sub> |

### 3.3 Quantification of 6-MN and possible effects of the amount present in the samples

Next, the amount of 6-MN was determined by UPLC-DAD for the different samples. The employed methodology, working in alkaline conditions, resulted in a chromatographic separation of the reference standard of nicotine (retention time of 6.5 minutes) and 6-MN (retention time of 7.7 minutes) as can be seen in the supplemental Figure S 20. All analyzed pouches showed a compound eluting at 7.7 minutes, but no peak was visible at 6.5 minutes. A linear relationship could be observed between 0.5 µg/mL and 20 µg/mL of 6-MN. Therefore, prior to each quantification, a mini-validation was performed accepting a recovery difference of 5% for an independently prepared reference standard for the concentration range of 5 µg/mL to 20 µg/mL.

The extract from the pouches was diluted with ammonium borate buffer at pH 9.0 until a concentration within this interval was achieved. The obtained quantification data for the different samples are presented in Table 3. The minimum and maximum average amounts found were 3.9 mg/bag and 19.4 mg/bag, corresponding to samples labeled Super Kick Extra Strong and Triple Kick Mega Strong, respectively. Upon comparison of the online labeled amount of 6-MN and the amount detected, no large discrepancies were detected as the samples contained a minimum of 78% and a maximum of 98% of the claimed amount. These results are in contrast with the very recent report on 6-NM containing e-liquids where only 12 to 13% of the claimed amount of 6-MN was found [26]. However, another recent study on e-liquids sold in Australia, demonstrated that quantities ranging from 2.9 to 100 mg/mL 6-MN were found, corresponding to 6.8 to 100% of the labeled amount, illustrating a large diversity in label accuracy [33].

**Table 3.** The amount of 6-MN claimed and the amount found. The data for the claimed amount was taken from the brand website (see supplemental data) as no clear quantity was mentioned on the sample itself. The quantification of each sample was performed on 3 separate pouches and each preparation was injected 4 times. These 12 injections were used to generate the mean amount of 6-MN present in the sample, while the minimum and maximum amount represent the mean amount present in one pouch of that particular sample. The measurement uncertainty was estimated as a confidence interval using the standard deviations obtained for these preparations. The measurement uncertainty (MUs) is expressed as a confidence interval using the standard deviation of the generated quantification results.

| **Strength and flavor** | **Claimed amount of 6-MN (mg/pouch)** | **Measured amount of 6-MN (mg/pouch ± MU)** | **Measured versus claimed 6-MN (%)** |
| --- | --- | --- | --- |
| Super kick - extra strong, double mint | 5 | 4.9 ± 0.2 | 98 |
| Super kick - extra strong, cola ice | 5 | 4.6 ± 0.2 | 92 |
| Super kick - extra strong, peach ice | 5 | 3.9 ± 0.2 | 78 |
| Double kick - super strong, ruby berry ice | 10 | 8.9 ± 0.4 | 89 |
| Double kick - super strong, menthol ice | 10 | 9.3 ± 0.3 | 93 |
| Double kick - super strong, muffin | 10 | 8.3 ± 0.6 | 83 |
| Triple kick - mega strong, mango ice | 20 | 19.4 ± 0.7 | 97 |
| Triple kick - mega strong, candy tobacco | 20 | 18.7 ± 0.6 | 94 |
| Triple kick - mega strong, muffin | 20 | 17.9 ± 0.7 | 90 |

## DISCUSSION

The analysis performed on these easily available “NoNIC” proclaimed, tobacco-free and nicotine-free; pouches demonstrated that these items were indeed devoid of nicotine, but contained the synthetic nicotine alkaloid 6-methyl nicotine (6-MN) and this to quantities ranging from 4 up to 20 mg of 6-MN per pouch representing respectively the lowest and highest “NoNIC” strengths. These values are quite similar to the quantities of nicotine found in pouches that were/are available in the United States of America [34] and Europe [35], which will likely result in pharmacological effects upon consumption. However, it should be noted that this amount represents the total amount in the pouches and does not reflect the amount which is released in the mouth upon consumption of the product nor the amount which is actually absorbed by the gastrointestinal system due to ingestion of the saliva. In contrast to inhalation from smoking, oromucosal and gastrointestinal absorption routes will result in a more gradual increase of nicotine concentrations in the brain and other peripheral tissues [36]. For this reason, it has been stated that oral nicotine pouches are less toxic and less addictive than cigarettes [37]. Nevertheless, although more than 500 mg of oral nicotine is likely required to cause a lethal overdose for an adult [38], severe acute oral nicotine intoxications, requiring medical assistance, have been reported for 1 mg in young children [39] or 10 mg in adults [40]. Based on the structural similarities with nicotine, it is likely that comparable oral concentrations of 6-MN might result in effects although one study reported that the molecule showed higher cytotoxicity levels than nicotine, but this was in human bronchial epithelial cell lines, representing the lung environment [41]. Moreover, pharmacological data on the binding potential of this molecule to the nicotine acetylcholine receptors are somewhat conflicting. One study in rats demonstrated up to 3.3 times higher affinity of 6-MN than nicotine, while another study, performed on mice, showed a lower binding affinity [ 42, 43]. Interestingly, functional activity studies testing the *in vivo* effect of 6-MN in rats or mice reported that this molecule was at least 2 to ± 5 times more potent than nicotine [42, 43]. It was postulated that the more lipophilic nature of 6-MN could result in enhanced membrane permeability and thus enhanced cellular uptake. Moreover, it was also found that the position of the methylation was also an important factor for nicotine binding to rat brain membranes, just like the size of the alkyl group. However, it remains to be elucidated if this is also the case for humans. Nevertheless, it’s evident from the lack of toxicological data and pharmacological data that much more in-depth *in vitro* or *in vivo* data is required to determine the precise action of this novel synthetic nicotinic alkaloid, taking into account the intake route. Yet, a part of a likely unaware/misled young adult subpopulation is exposed to this molecule through these pouches. Some European countries, such as the Netherlands, that previously classified nicotine pouches as food products, can easily ban these 6-MN pouches based on toxicological extrapolation of existing data on nicotine [27]. However, these items remain unregulated in many other European countries, and consequently due to the trade agreement amongst EU members and also with former Member States, part of a likely unaware/misled young adult subpopulation has easy access to these items. Especially considering that this molecule will likely, similarly to nicotine, have detrimental effects on the still-developing brain of youngsters and young adults, the target population of these pouches. Therefore, it is pivotal for the different Member States to decide how to classify these pouches , either as food products, tobacco products, or other, and to strive for some sort of uniformization to better protect the health of its young citizens.

## ACKNOWLEDGEMENTS

We kindly would like to thank the inspectors of the Belgian Federal Public Service Health, Food Chain Safety and Environment, responsible for the control of tobacco and related products and Sciensano’s chromatography platform for the constructive collaboration.

## AUTHOR CONTRIBUTIONS

C.V. and S.B conceptualized and designed the study. C.V, M.D, M.C. and E.T. carried out the chemical analysis. C.V. and E.T. wrote the first draft and S.B., M.D. and M.C. and E.T. contributed to the revision of the manuscript.

## DATA AVAILABILITY

The data supporting this study’s findings are available in this article’s supplementary material.

## REFERENCES

[1] Bainton RJ, Tsai LT, Singh CM, Moore MS, Neckameyer WS, Heberlein U. Dopamine modulates acute responses to cocaine, nicotine and ethanol in Drosophila. Curr Biol. 2000;10(4):187–194. doi:10.1016/s0960-9822(00)00336-5.

[2] Benowitz NL. Pharmacology of nicotine: addiction, smoking-induced disease, and therapeutics. Annu Rev Pharmacol Toxicol. 2009;49:57–71. doi:10.1146/annurev.pharmtox.48.113006.094742

[3] Mao D, Gallagher K, McGehee DS. Nicotine potentiation of excitatory inputs to ventral tegmental area dopamine neurons. J Neurosci. 2011 May 4;31(18):6710–20. doi: 10.1523/JNEUROSCI.5671-10.2011.

[4] Wei C, Han X, Weng D, Feng Q, Qi X, Li J, Luo M. Response dynamics of midbrain dopamine neurons and serotonin neurons to heroin, nicotine, cocaine, and MDMA. Cell Discov. 2018 Nov 6;4:60. doi: 10.1038/s41421-018-0060-z.

[5] Wolter M, Lapointe T, Melanson B, et al. Memory enhancing effects of nicotine, cocaine, and their conditioned stimuli; effects of beta-adrenergic and dopamine D2 receptor antagonists. Psychopharmacology (Berl). 2021;238(9):2617–2628. doi:10.1007/s00213-021-05884-x

[6] Valentine G, Sofuoglu M. Cognitive Effects of Nicotine: Recent Progress. Curr Neuropharmacol. 2018;16(4):403–414. doi:10.2174/1570159X15666171103152136

[7] Beer A. Chapter 27 - Nicotine and Cognition: Effects of Nicotine on Attention and Memory Systems in Humans, Neuropathology of Drug Addictions and Substance Misuse, Academic Press, 2016, ISBN: 9780128002131. 10.1016/B978-0-12-800213-1.00027-4

[8] Seoane-Collazo P, Diéguez C, Nogueiras R, Rahmouni K, Fernández-Real JM, López M. Nicotine’ actions on energy balance: Friend or foe?. Pharmacol Ther. 2021;219:107693. doi:10.1016/j.pharmthera.2020.107693

[9] Duke, D., Wohlgemuth, E., Adams, K.R. et al. Earliest evidence for human use of tobacco in the Pleistocene Americas. Nat Hum Behav 6, 183–192 (2022). 10.1038/s41562-021-01202-9

[10] Proctor RN. The history of the discovery of the cigarette-lung cancer link: evidentiary traditions, corporate denial, global toll. Tob Control. 2012;21(2):87–91. doi:10.1136/tobaccocontrol-2011-050338

[11] Schmidt BL, Dierks EJ, Homer L, Potter B. Tobacco smoking history and presentation of oral squamous cell carcinoma. J Oral Maxillofac Surg. 2004 Sep;62(9):1055–8. doi: 10.1016/j.joms.2004.03.010..

[12] Dai X, Gakidou E, Lopez AD. Evolution of the global smoking epidemic over the past half century: strengthening the evidence base for policy action. Tob Control. 2022;31(2):129–137. doi:10.1136/tobaccocontrol-2021-056535

[13] European commission, Directive 2001/37/EC of the European Parliament and of the Council of 5 June 2001 on the approximation of the laws, regulations and administrative provisions of the Member States concerning the manufacture, presentation and sale of tobacco products - Commission statement. https://eur-lex.europa.eu/legal-content/en/ALL/?uri=CELEX%3A32001L0037 [Accessed on 13/09/2024]

[14] Bertollini R, Ribeiro S, Mauer-Stender K, Galea G. Tobacco control in Europe: a policy review. Eur Respir Rev. 2016;25(140):151–157. doi:10.1183/16000617.0021-2016

[15] European commission. Directive 2014/40/EU of the European Parliament and of the Council of 3 April 2014 on the approximation of the laws, regulations and administrative provisions of the Member States concerning the manufacture, presentation and sale of tobacco and related products and repealing Directive 2001/37/EC Text with EEA relevance. http://data.europa.eu/eli/dir/2014/40/oj [Accessed on 13/09/2024].

[16] Vardavas CI. European Tobacco Products Directive (TPD): current impact and future steps. Tob Control. 2022;31(2):198–201. doi:10.1136/tobaccocontrol-2021-056548

[17] Duren M, Atella L, Welding K, Kennedy RD. Nicotine pouches: a summary of regulatory approaches across 67 countries. Tob Control. 2023 Feb 7:tc-2022-057734. doi: 10.1136/tc-2022-057734.

[18] M. Jackson, J., Weke, A. & Holliday, R. Nicotine pouches: a review for the dental team. Br Dent J 235, 643–646 (2023). doi:10.1038/s41415-023-6383-7

[19] The German Federal Institute for Risk Assessment, Bfr. https://mobil.bfr.bund.de/cm/349/health-risk-assessment-of-nicotine-pouches.pdf [accessed 13/09/2024]

[20] Agence nationale de sécurité sanitaire de l’alimentation, https://www.anses.fr/fr/system/files/Toxicovigilance2023AUTO0121Ra.pdf [accessed 13/09/2024]

[21] Mallock N, Schulz T, Malke S, Dreiack N, Laux P, Luch A. Levels of nicotine and tobacco-specific nitrosamines in oral nicotine pouches. Tob Control. 2024 Feb 20;33(2):193–199. doi: 10.1136/tc-2022-057280.

[22] Castro EM, Lotfipour S, Leslie FM. Nicotine on the developing brain. Pharmacol Res. 2023;190:106716. doi:10.1016/j.phrs.2023.106716

[23] Keogh, A. Nicotine pouches. Br Dent J 230, 61–62 (2021). 10.1038/s41415-021-2622-y

[24] Belgian government, Royal decree of 14^th^ March 2024, https://etaamb.openjustice.be/nl/koninklijk-besluit-van-14-maart-2023_n2023041247.html <u>[accessed 13/09/2024]</u>

[25] Jordt SE, Jabba SV, Zettler PJ, Berman ML. Spree Bar, a vaping system delivering a synthetic nicotine analogue, marketed in the USA as ‘PMTA exempt’. Tob Control. 2024 Mar 18:tc-2023-058469. doi: 10.1136/tc-2023-058469.

[26] Erythropel HC, Jabba SV, Silinski P, Anastas PT, Krishnan-Sarin S, Zimmerman JB, Jordt SE. Variability in Constituents of E-Cigarette Products Containing Nicotine Analogues. JAMA. 2024 Aug 7:e2412408. doi: 10.1001/jama.2024.12408. Epub ahead of print.

[27] Dutch National Institute for Public Health and the Environment, RIVM. https://www.rivm.nl/documenten/beoordeling-van-6-methylnicotinegehalte-in-nicotinezakjes [accessed 13/09/2024]

[28] Vanhee C, Jacobs B, Mori M, Kamugisha A, Debehault L, Canfyn M, Ceyssens B, Van Der Meersch H, van Hoorde K, Deconinck E, et al. Uncovering the Quality Deficiencies with Potentially Harmful Effects in Substandard and Falsified PDE-5 Inhibitors Seized by Belgian Controlling Agencies. Forensic Sciences. 2023; 3(3):426–451. 10.3390/forensicsci3030031

[29] Barhdadi S, Desmedt B, Courselle P, Rogiers V, Vanhaecke T, Deconinck E. A simple dilute-and-shoot method for screening and simultaneous quantification of nicotine and alkaloid impurities in electronic cigarette refills (e-liquids) by UHPLC-DAD. J Pharm Biomed Anal. 2019 May 30;169:225–234. doi: 10.1016/j.jpba.2019.03.002.

[30] Digard H, Gale N, Errington G, Peters N, McAdam K. Multi-analyte approach for determining the extraction of tobacco constituents from pouched snus by consumers during use. Chem Cent J. 2013;7(1):55. Published 2013 Apr 2. doi:10.1186/1752-153X-7-55

[31] Haglid F. The Methylation of Nicotine with Methyl-litium. Acta Chem Scand. 1967; 21:329–334. 10.3891/acta.chem.scand.21-0329

[32] Seeman JI, Secor HV, Chavdarian CG, Sanders EB, Bassfield RL, Whidby JF. Steric and Conformational Effects in Nicotine Chemistry. J Org Chem. 1981; 46:3040–3048. 10.1021/jo00328a010

[33] Jenkins C, Kelso C, Morgan J. 6-Methylnicotine: a new nicotine alternative identified in e-cigarette liquids sold in Australia. Med J Aust. 2024. doi: 10.5694/mja2.52423.

[34] Stanfill S, Tran H, Tyx R, Fernandez C, Zhu W, Marynak K, King B, Valentín-Blasini L, Blount BC, Watson C. Characterization of Total and Unprotonated (Free) Nicotine Content of Nicotine Pouch Products. Nicotine Tob Res. 2021 Aug 18;23(9):1590–1596. doi: 10.1093/ntr/ntab030.

[35] Mallock N, Schulz T, Malke S, Dreiack N, Laux P, Luch A. Levels of nicotine and tobacco-specific nitrosamines in oral nicotine pouches. Tob Control. 2024 Feb 20;33(2):193–199. doi: 10.1136/tc-2022-057280.

[36] Kolli AR, Calvino-Martin F, Kuczaj AK, Wong ET, Titz B, Xiang Y, Lebrun S, Schlage WK, Vanscheeuwijck P, Hoeng J. Deconvolution of Systemic Pharmacokinetics Predicts Inhaled Aerosol Dosimetry of Nicotine. Eur J Pharm Sci. 2023; 180:106321. doi: 10.1016/j.ejps.2022.106321.

[37] McEwan, M., Azzopardi, D., Gale, N., Camacho, O. M., Hardie, G., Fearon, I. M., & Murphy, J. (2022). A Randomised Study to Investigate the Nicotine Pharmacokinetics of Oral Nicotine Pouches and a Combustible Cigarette. Eur J Drug Metab Pharmacokinet. 2022;47:211–221. doi:10.1007/s13318-021-00742-9

[38] Mayer B. How much nicotine kills a human? Tracing back the generally accepted lethal dose to dubious self-experiments in the nineteenth century. Arch Toxicol. 2014;88(1):5–7. doi:10.1007/s00204-013-1127-0

[39] Chang JT, Wang B, Chang CM, Ambrose BK. National estimates of poisoning events related to liquid nicotine in young children treated in US hospital emergency departments, 2013-2017. Inj Epidemiol. 2019;6:10. doi:10.1186/s40621-019-0188-9

[40] Cho YS, Sohn Y. Acute heart failure after oral intake of liquid nitoctine. Soonchunhyang Med Sci 2020; 26(1): 22–25. doi: doi.org/10.15746/sms.20.006

[41] Qi H, Chang X, Wang K, Xu Q, Liu M, Han B. Comparative analyses of transcriptome sequencing and carcinogenic exposure toxicity of nicotine and 6-methyl nicotine in human bronchial epithelial cells. Toxicol In Vitro. 2023;93:105661. doi:10.1016/j.tiv.2023.105661

[42] Wang DX, Booth H, Lerner-Marmarosh N, Osdene TS, Abood LG. Structure–activity relationships for nicotine analogs comparing competition for [3H]nicotine binding and psychotropic potency. Drug development research. 1998; 45; 1. doi.org/10.1002/(SICI)1098-2299(199809)45:1<10::AID-DDR2>3.0.CO;2-G

[43] Dukat M, El-Zahabi M, Ferretti G, Damaj MI, Martin BR, Young R, Glennon RA. (-)6-n-Propylnicotine antagonizes the antinociceptive effects of (-)nicotine. Bioorg Med Chem Lett. 2002 Oct 21;12(20):3005–7. doi: 10.1016/s0960-894x(02)00614-5.

