## supplemental figure S1 for "The emergence of a novel synthetic nicotine analog 6-methyl nicotine (6-MN) in proclaimed tobacco- and nicotine-free pouches available in Europe"

A

### Aroma King Nicotine Pouches Triple Kick

Experience the powerful kick of Aroma King Nicotine Pouches Triple Kick. These premium pouches provide you with a **bold** and **satisfying** nicotine hit without any **tobacco**. Enjoy the convenience and discretion of these pouches, perfect for on-the-go use. Unleash the triple kick of flavour, nicotine, and portability with Aroma King.

**Product Features:**

Nicotine Strength: 20mg
Manufacturer: Aroma King
Format: Slim
Weight 0.5gr per pouch
Contains 25 Pouches
**Does Not Contain Tobacco & Nicotine**

B

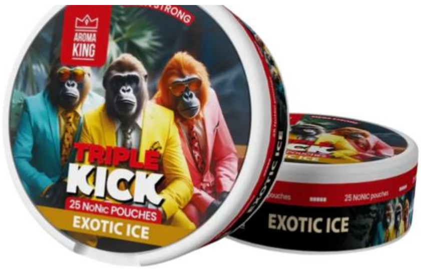


− 1 +

Add to cart

Buy it now

Aroma King NoNic Triple Kick Exotic Ice nicotine pouches come from the Polish brand Aroma King. These pouches pack an amazing taste of exotic fruits. It's a must try nicotine pouch for those who are seeking a flavorful kick.

The pouches strength are 20 mg and packs a real punch. The Aroma King range is mainly used by more experienced nicotine pouch users.

- **Brand Name: Aroma King**
- **Product Type: Nicotine Pouches**
- **Pouch Format: Slim**
- **Our Strength Rating: Strong**
- **Nicotine Per Pouch: 20 mg/g**
- **Flavour: Exotic Ice**
- **Pouches Per Can: 25**

C

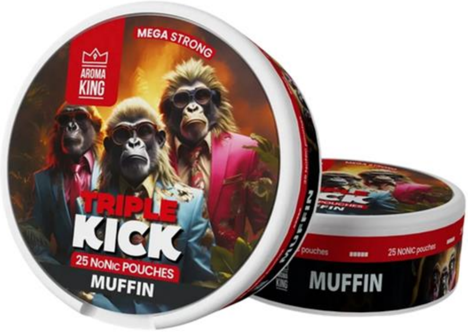


Aroma King NoNic® Triple Kick Muffin - 20mg

£ 4.99

Quantity

1

Add To Cart

Buy Now

- Flavour - **Muffin**
- Portion size - **Slim**
- Strength - **Extra strong**
- Pouches per can - **25**
- NoNicotine® Level - **20mg**
- Comparative nicotine level - **100mg**
- Net weight - **16 g**
- Product type - **NoNicotine® pouches**

NoNic® is a brand new alternative to traditional nicotine. NoNic is a non-toxic, non-addictive and non-psychoactive substance, and has an effect just like nicotine.

1mg of NoNic® is equivalent to 5mg of Nicotine !
