## Supplemental figure S2-S19 for "The emergence of a novel synthetic nicotine analog 6-methyl nicotine (6-MN) in proclaimed tobacco- and nicotine-free pouches available in Europe"

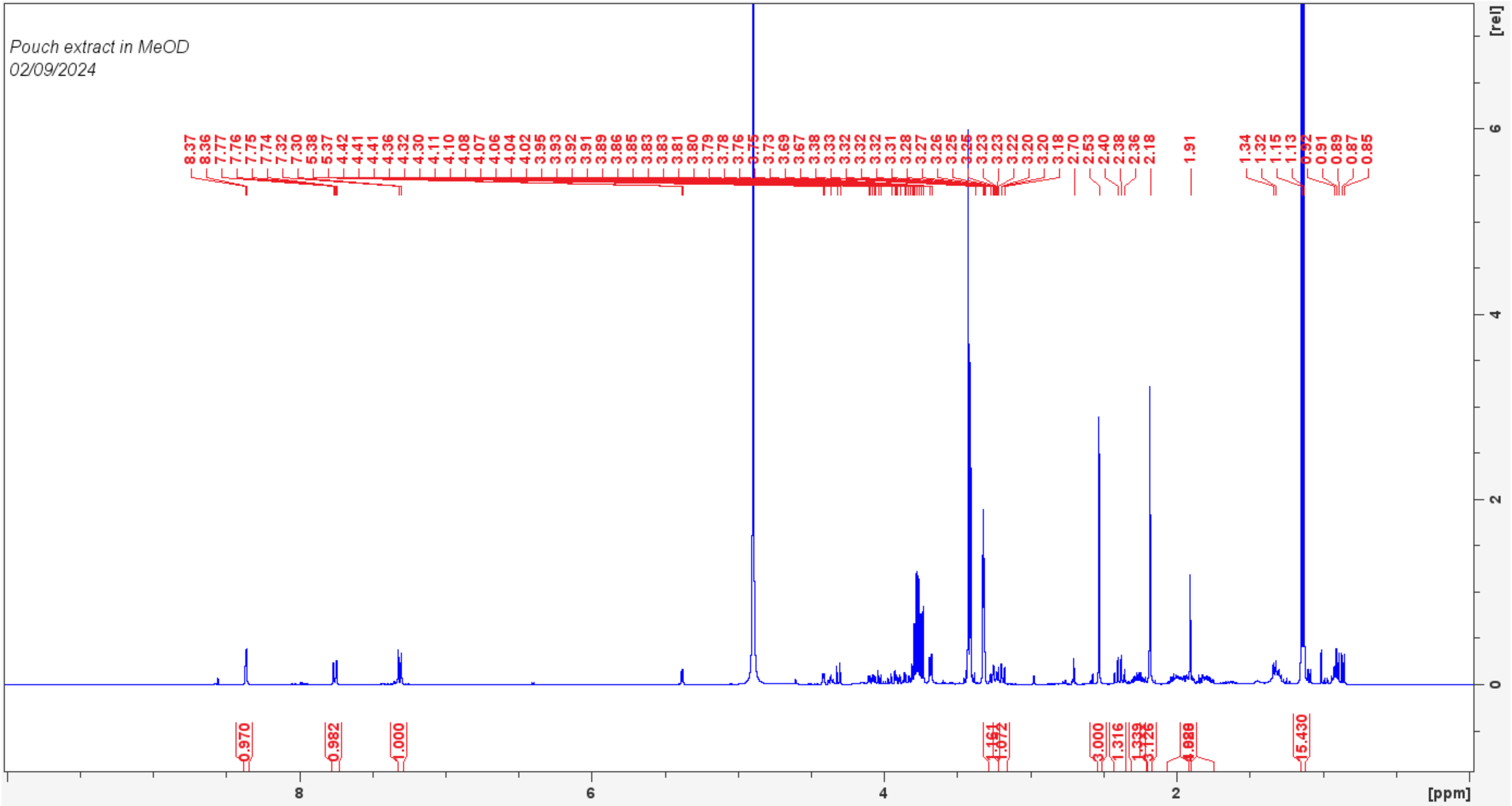

Pouch extract in MeOD
02/09/2024
8.37 8.36 7.77 7.76 7.75 7.74 7.32 7.30 5.38 5.37 4.42 4.41 4.41 4.36 4.32 4.30 4.11 4.10 4.08 4.07 4.06 4.04 4.02 3.95 3.93 3.92 3.91 3.89 3.86 3.85 3.83 3.83 3.81 3.80 3.79 3.78 3.76 3.75 3.73 3.69 3.67 3.38 3.33 3.32 3.32 3.32 3.31 3.28 3.27 3.26 3.25 3.25 3.23 3.23 3.22 3.20 3.20 3.18 2.70 2.53 2.40 2.38 2.36 2.18 1.91 1.34 1.32 1.15 1.13 0.92 0.91 0.89 0.87 0.85
0.970
0.982
1.000
1.072
3.000
1.316
3.126
15.430
[rel]
6
4
2
0
8
6
4
2
[ppm]

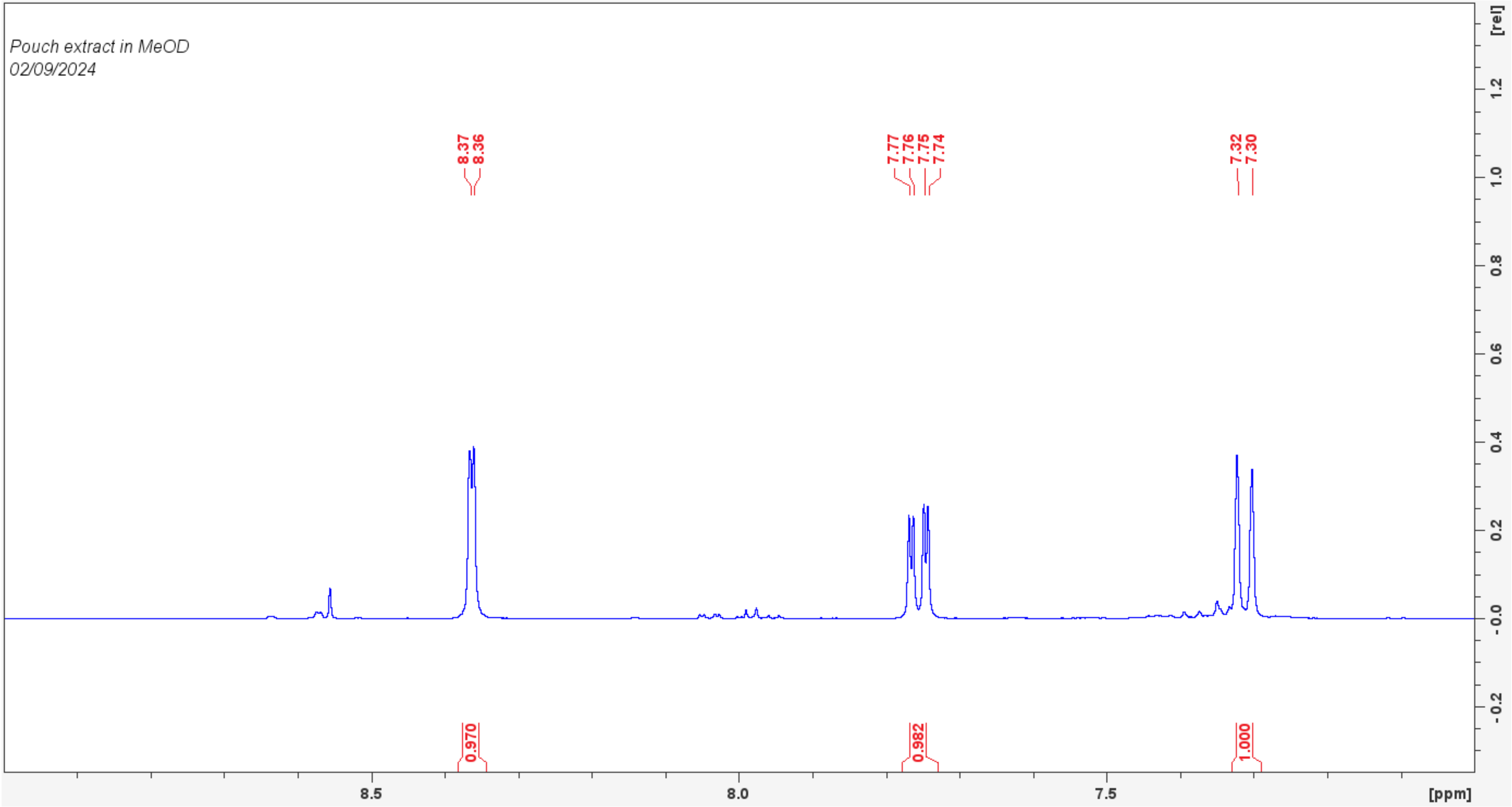

Pouch extract in MeOD
02/09/2024
8.37
8.36
7.77
7.76
7.75
7.74
7.32
7.30
0.970
0.982
1.000
[rel]
1.2
1.0
0.8
0.6
0.4
0.2
0.0
-0.2
8.5
8.0
7.5
[ppm]

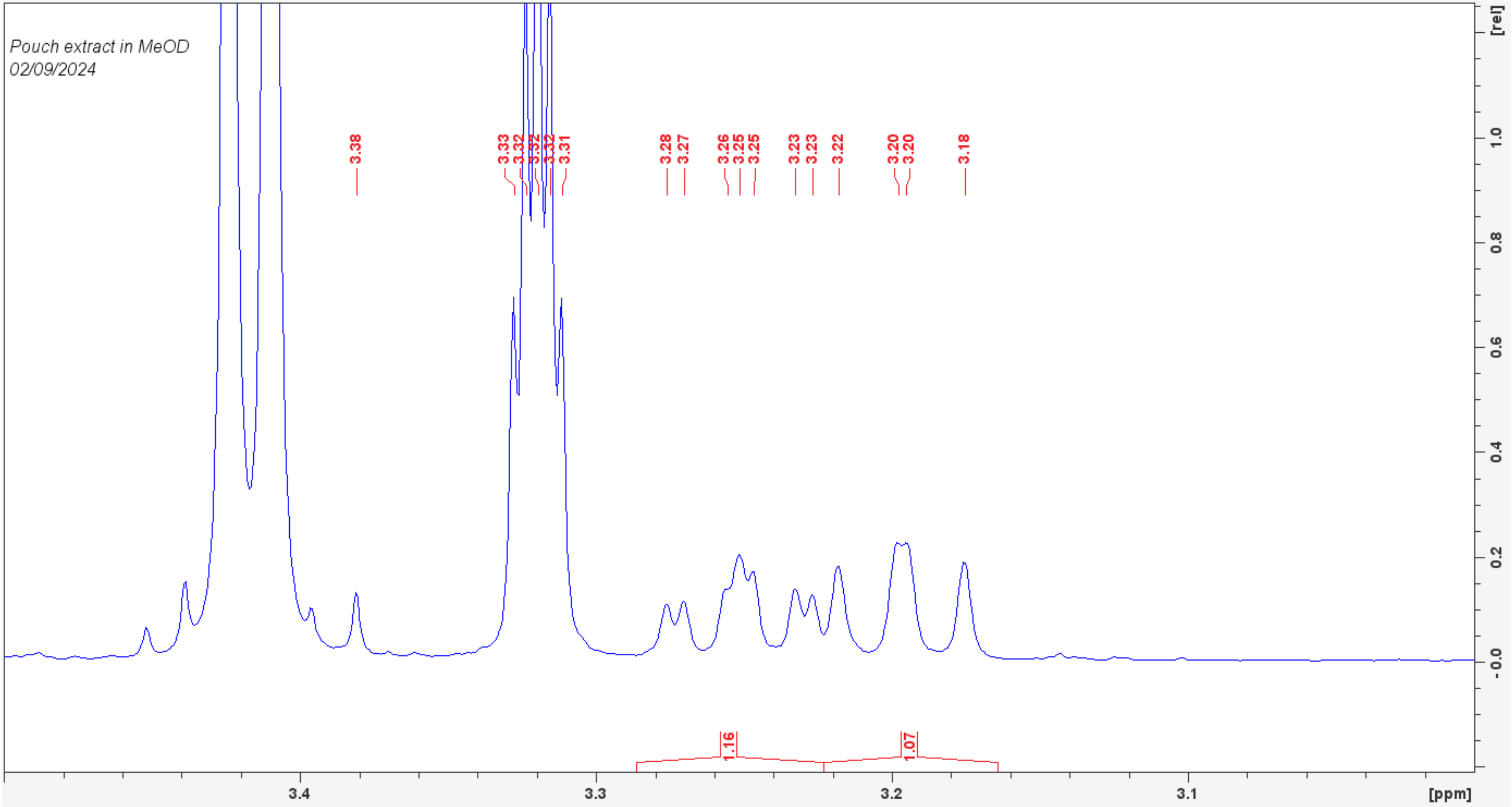

Pouch extract in MeOD
02/09/2024
3.38
3.33
3.32
3.32
3.32
3.31
3.28
3.27
3.26
3.25
3.25
3.23
3.23
3.22
3.20
3.20
3.18
1.16
1.07
3.4
3.3
3.2
3.1
[ppm]
[rel]
1.0
0.8
0.6
0.4
0.2
-0.0

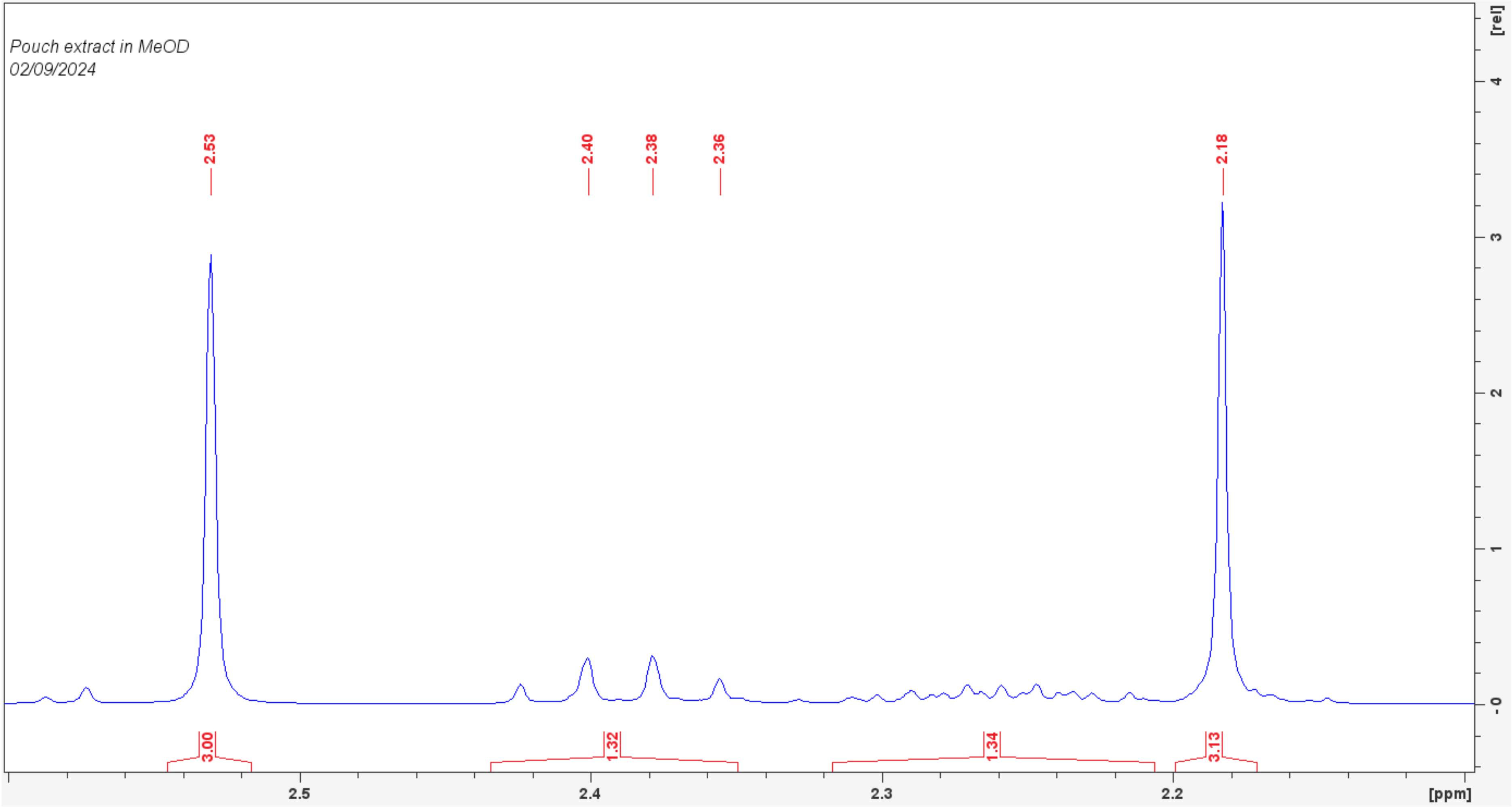
Pouch extract in MeOD
02/09/2024
2.53
2.40
2.38
2.36
2.18
3.00
1.32
1.34
3.13
2.5
2.4
2.3
2.2
[ppm]
[rel]
4
3
2
1
0

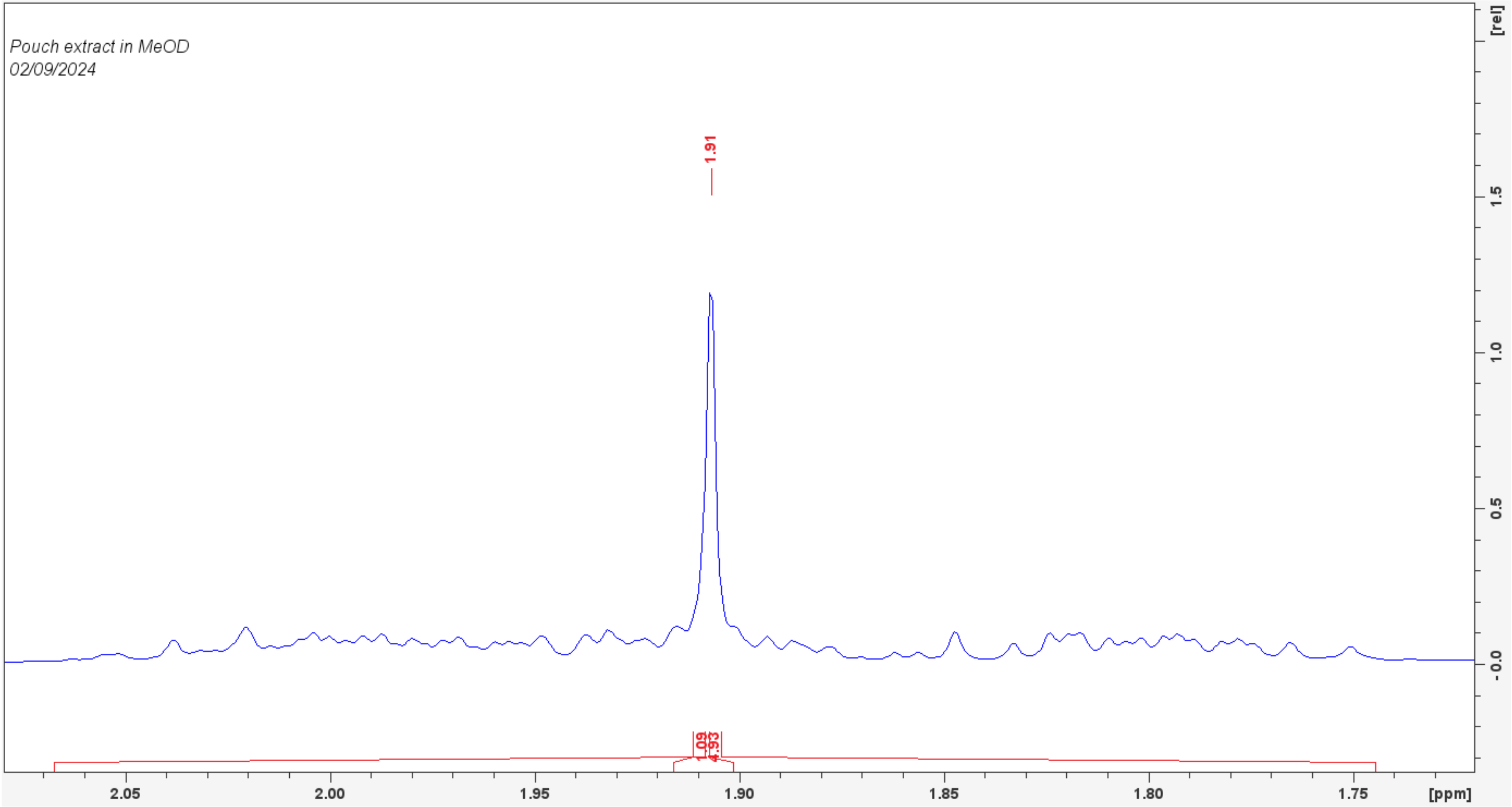

Pouch extract in MeOD
02/09/2024
1.91
1.09
4.93
2.05
2.00
1.95
1.90
1.85
1.80
1.75
[ppm]
[rel]
1.5
1.0
0.5
-0.0

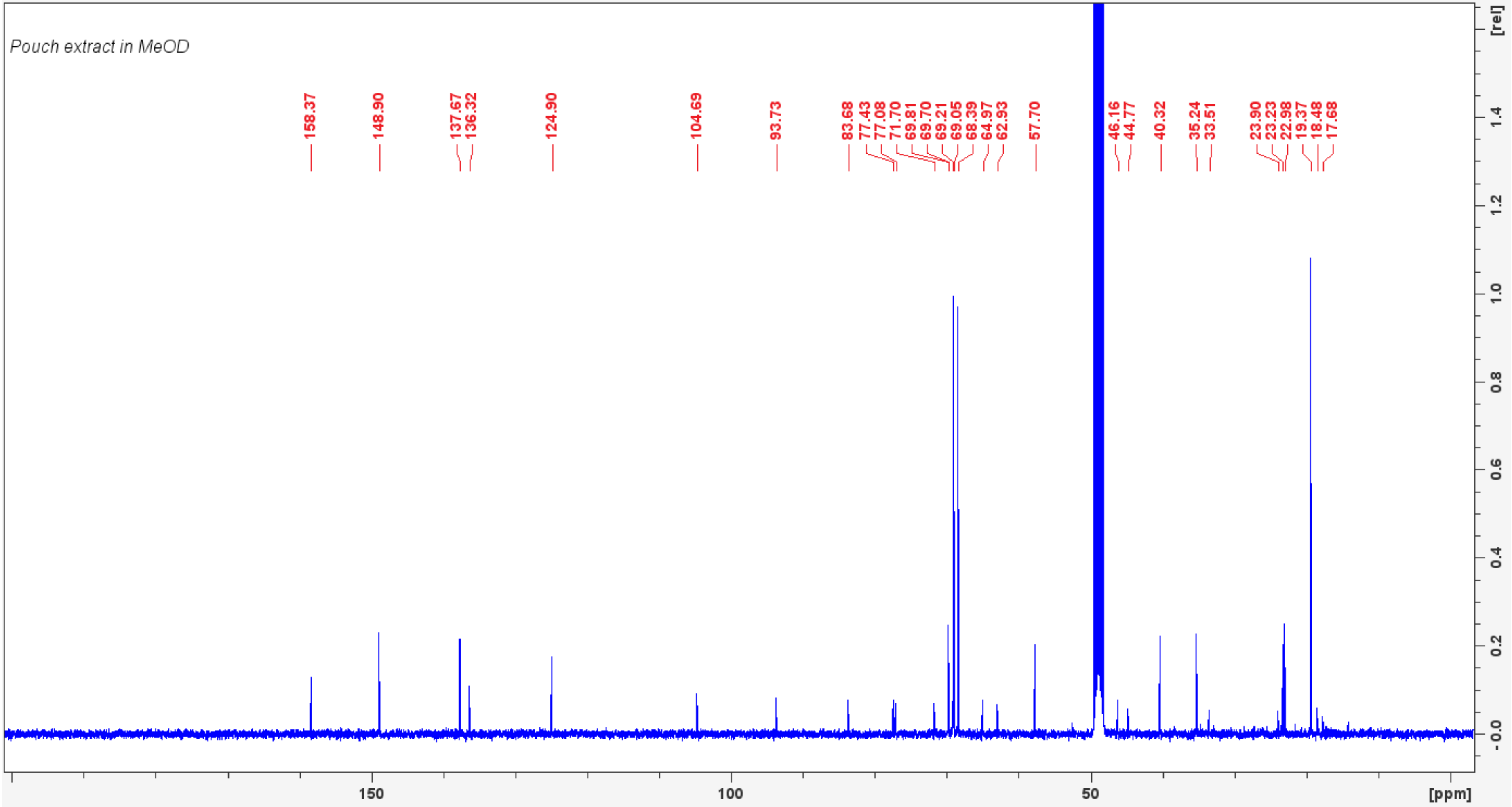
Pouch extract in MeOD
158.37
148.90
137.67
136.32
124.90
104.69
93.73
83.68
77.43
77.08
71.70
69.81
69.70
69.21
69.05
68.39
64.97
62.93
57.70
46.16
44.77
40.32
35.24
33.51
23.90
23.23
22.98
19.37
18.48
17.68
[rel]
1.4
1.2
1.0
0.8
0.6
0.4
0.2
-0.0
150
100
50
[ppm]

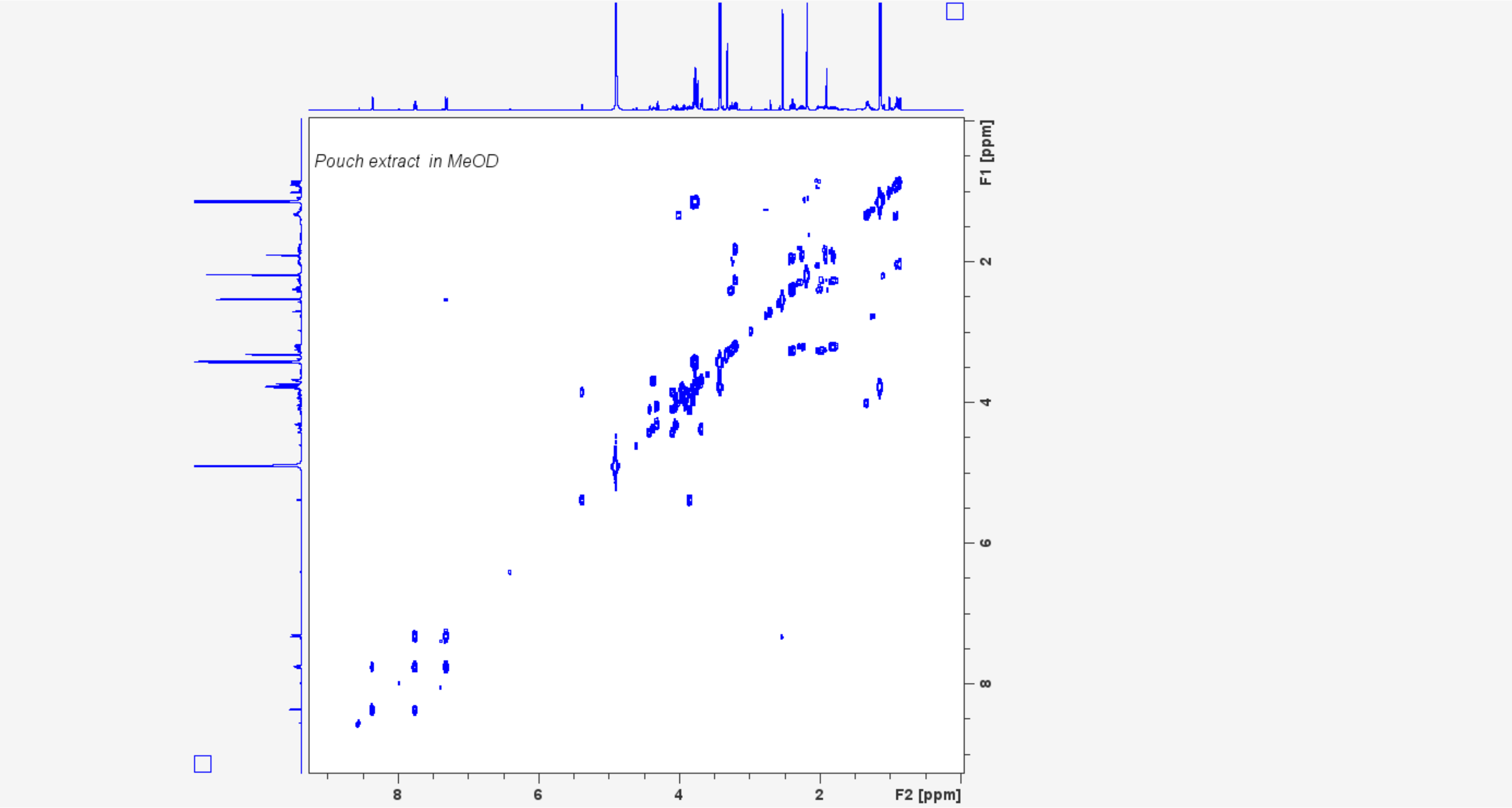

Pouch extract in MeOD
F1 [ppm]
F2 [ppm]
8
6
4
2

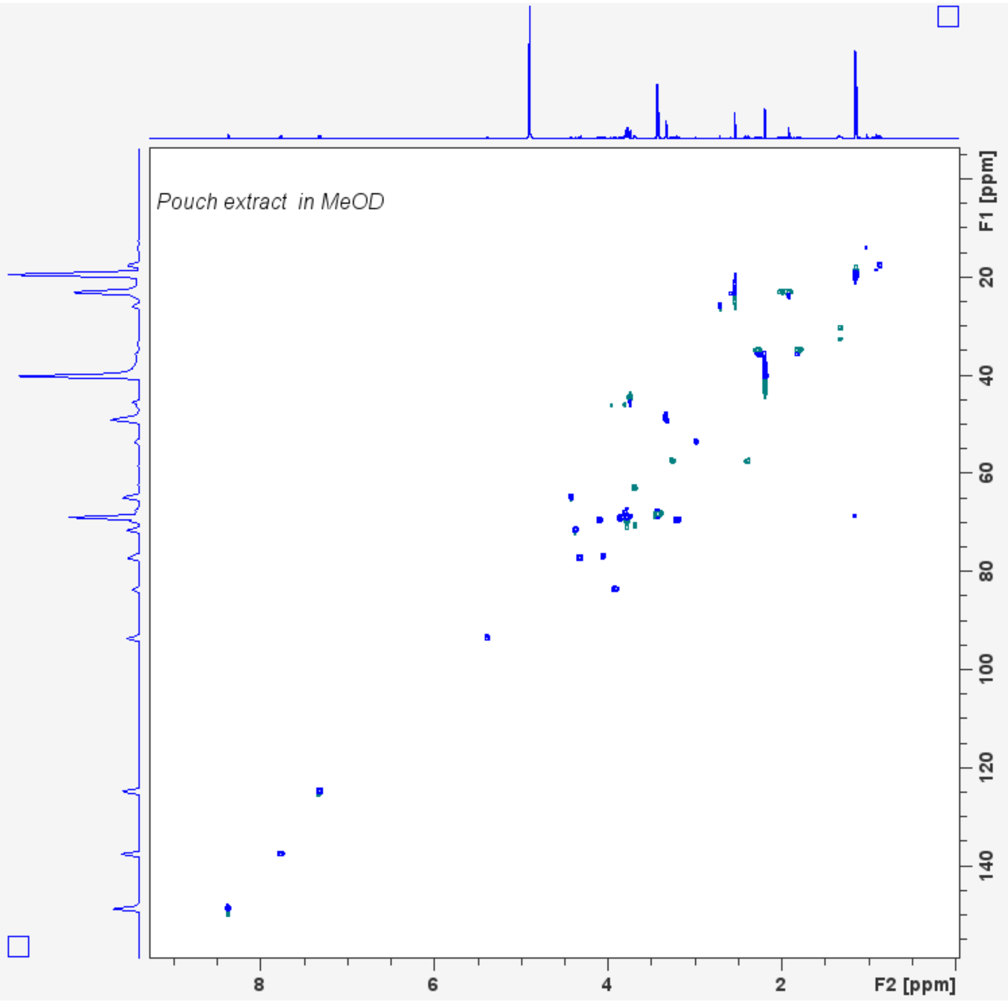

Pouch extract in MeOD
F1 [ppm]
20
40
60
80
100
120
140
8
6
4
2
F2 [ppm]

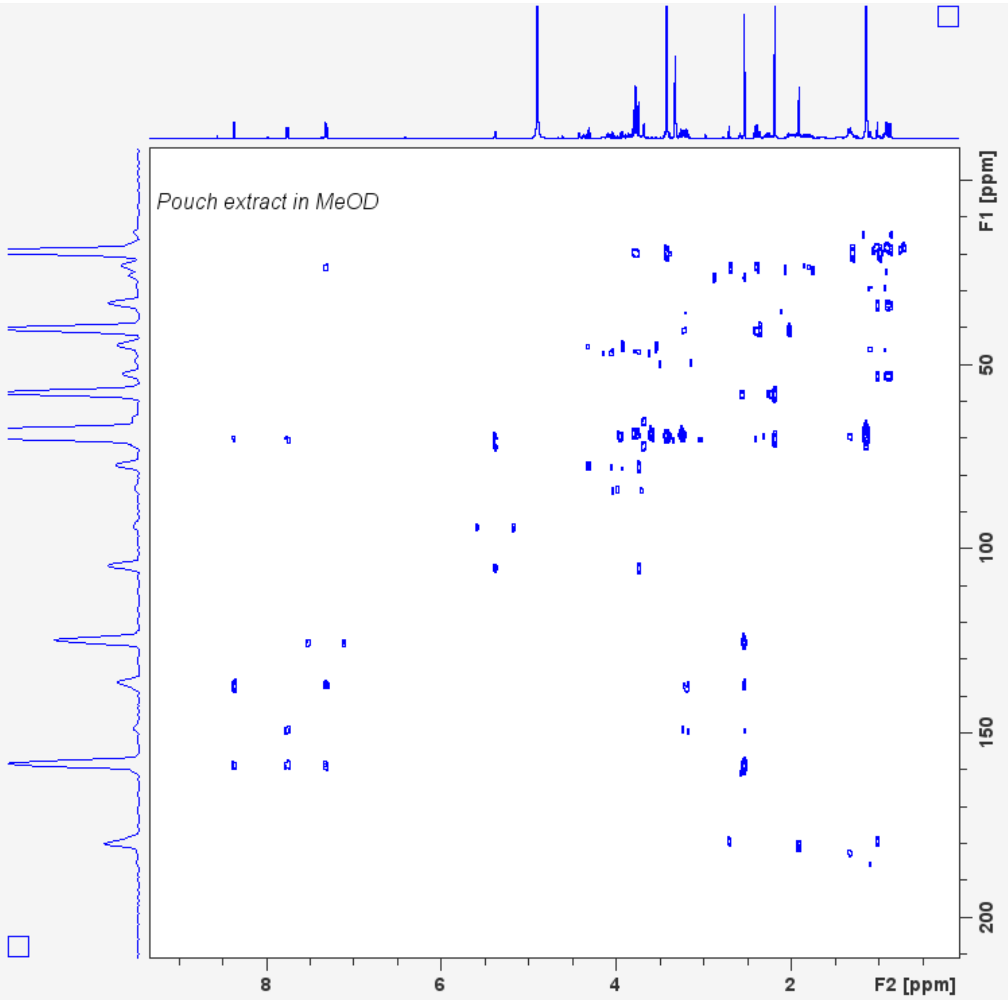

Pouch extract in MeOD
F1 [ppm]
50
100
150
200
8
6
4
2
F2 [ppm]

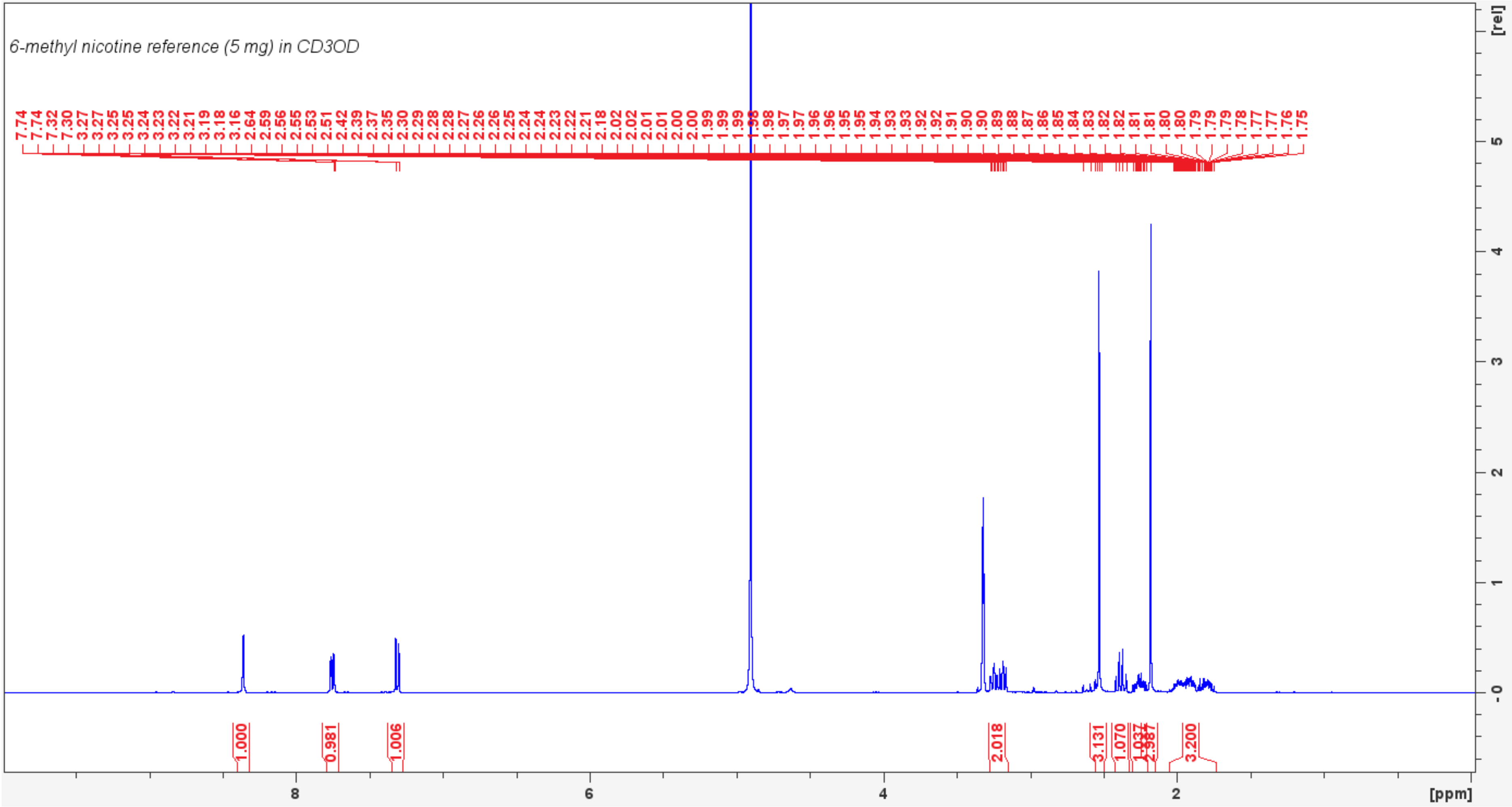
6-methyl nicotine reference (5 mg) in CD3OD
7.74 7.74 7.32 7.30 3.27 3.27 3.25 3.25 3.24 3.23 3.22 3.21 3.19 3.18 3.16 2.64 2.59 2.56 2.55 2.53 2.51 2.42 2.39 2.37 2.35 2.30 2.29 2.28 2.28 2.27 2.26 2.25 2.24 2.24 2.23 2.22 2.21 2.18 2.02 2.02 2.01 2.01 2.00 2.00 1.99 1.99 1.99 1.98 1.98 1.97 1.97 1.96 1.96 1.95 1.95 1.94 1.93 1.93 1.92 1.92 1.91 1.90 1.90 1.89 1.88 1.87 1.86 1.85 1.84 1.83 1.82 1.82 1.81 1.81 1.80 1.80 1.79 1.79 1.79 1.78 1.77 1.77 1.76 1.75
1.000
0.981
1.006
2.018
3.131
1.070
1.037
2.987
3.200
8
6
4
2
[ppm]
[rel]
5
4
3
2
1
0

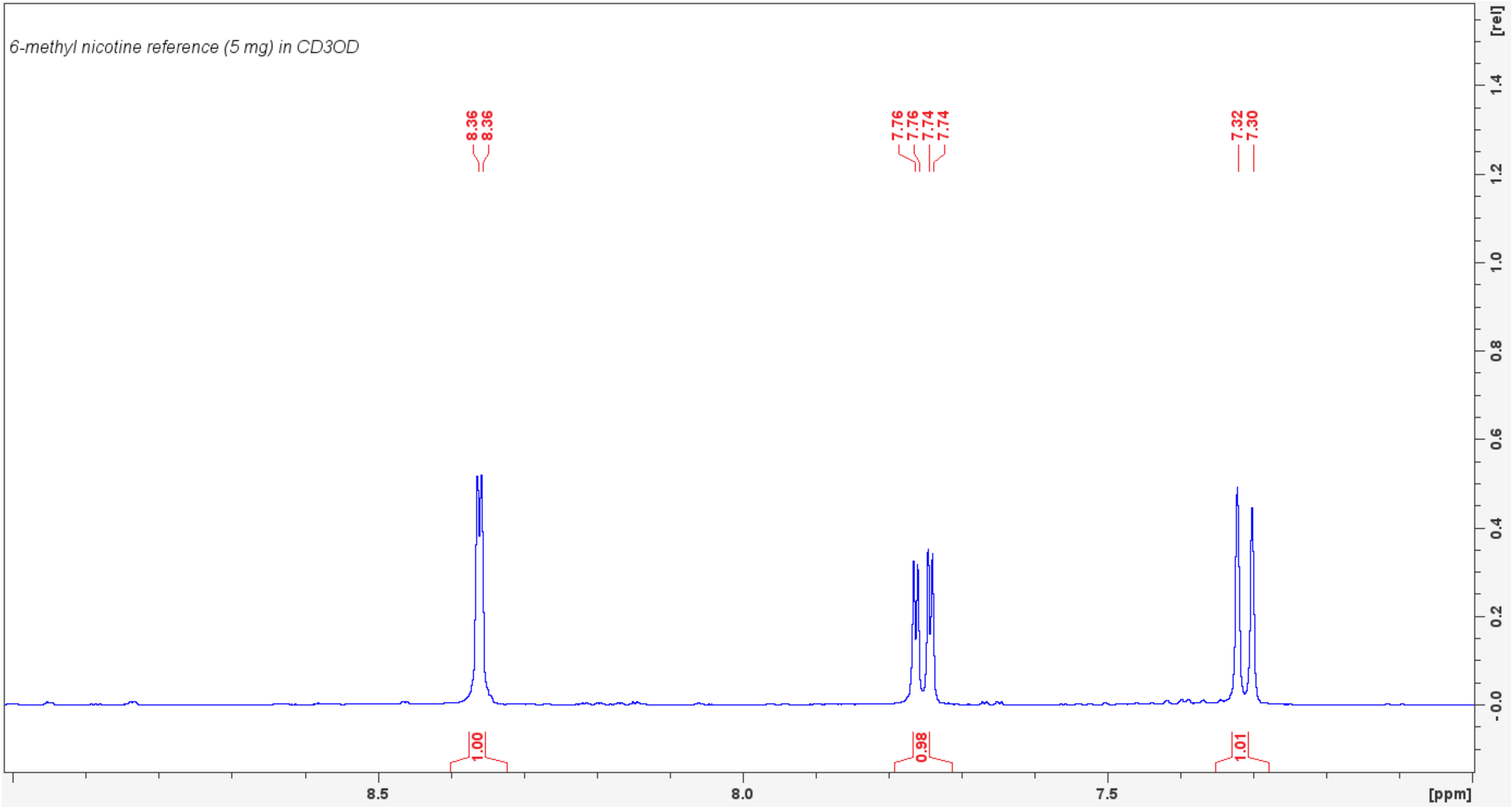

6-methyl nicotine reference (5 mg) in CD3OD
8.36
8.36
7.76
7.76
7.74
7.74
7.32
7.30
1.00
0.98
1.01
8.5
8.0
7.5
[ppm]
[rel]
1.4
1.2
1.0
0.8
0.6
0.4
0.2
-0.0

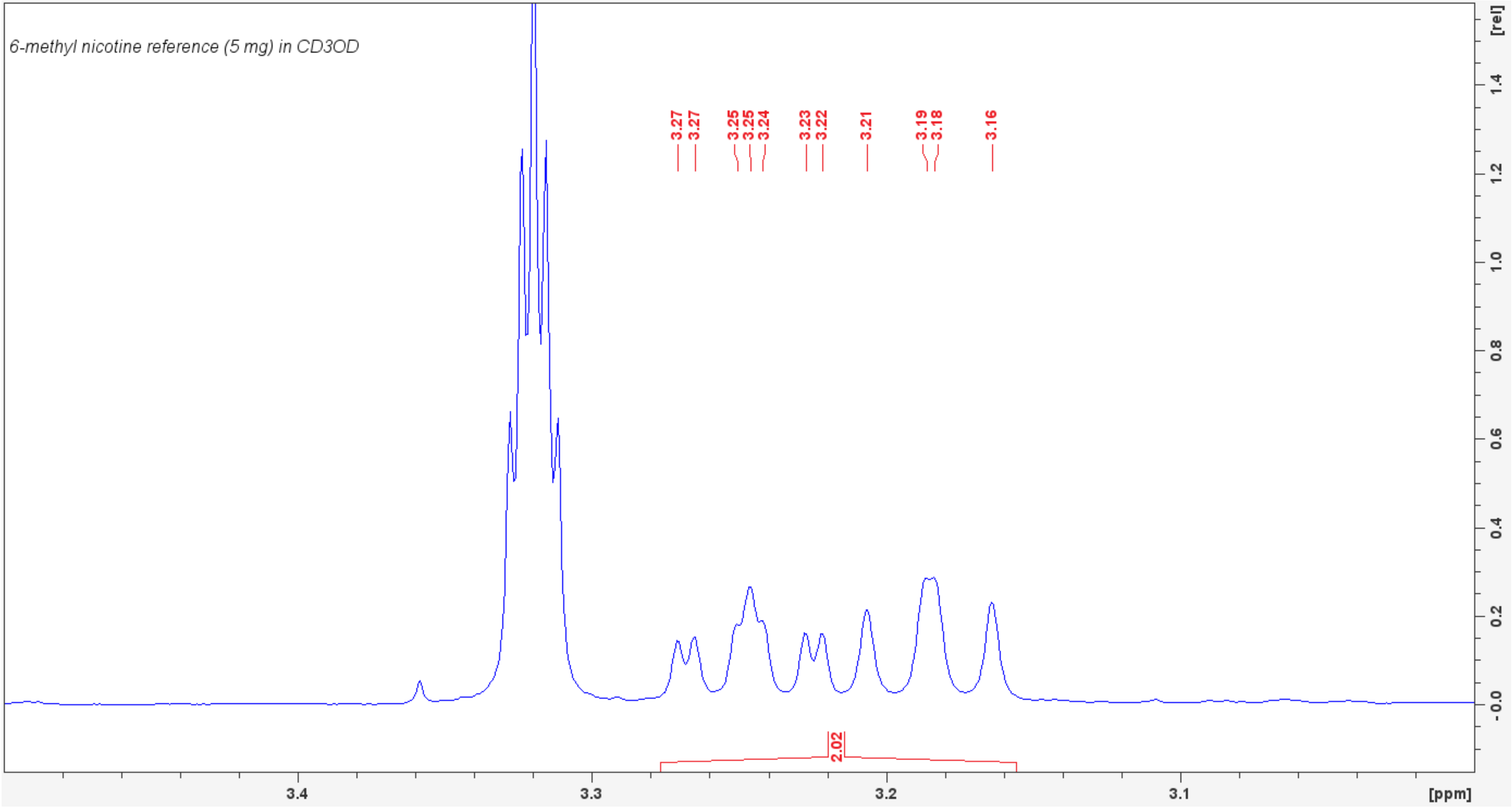

6-methyl nicotine reference (5 mg) in CD3OD
3.27
3.27
3.25
3.25
3.24
3.23
3.22
3.21
3.19
3.18
3.16
2.02
3.4
3.3
3.2
3.1
[ppm]
[rel]

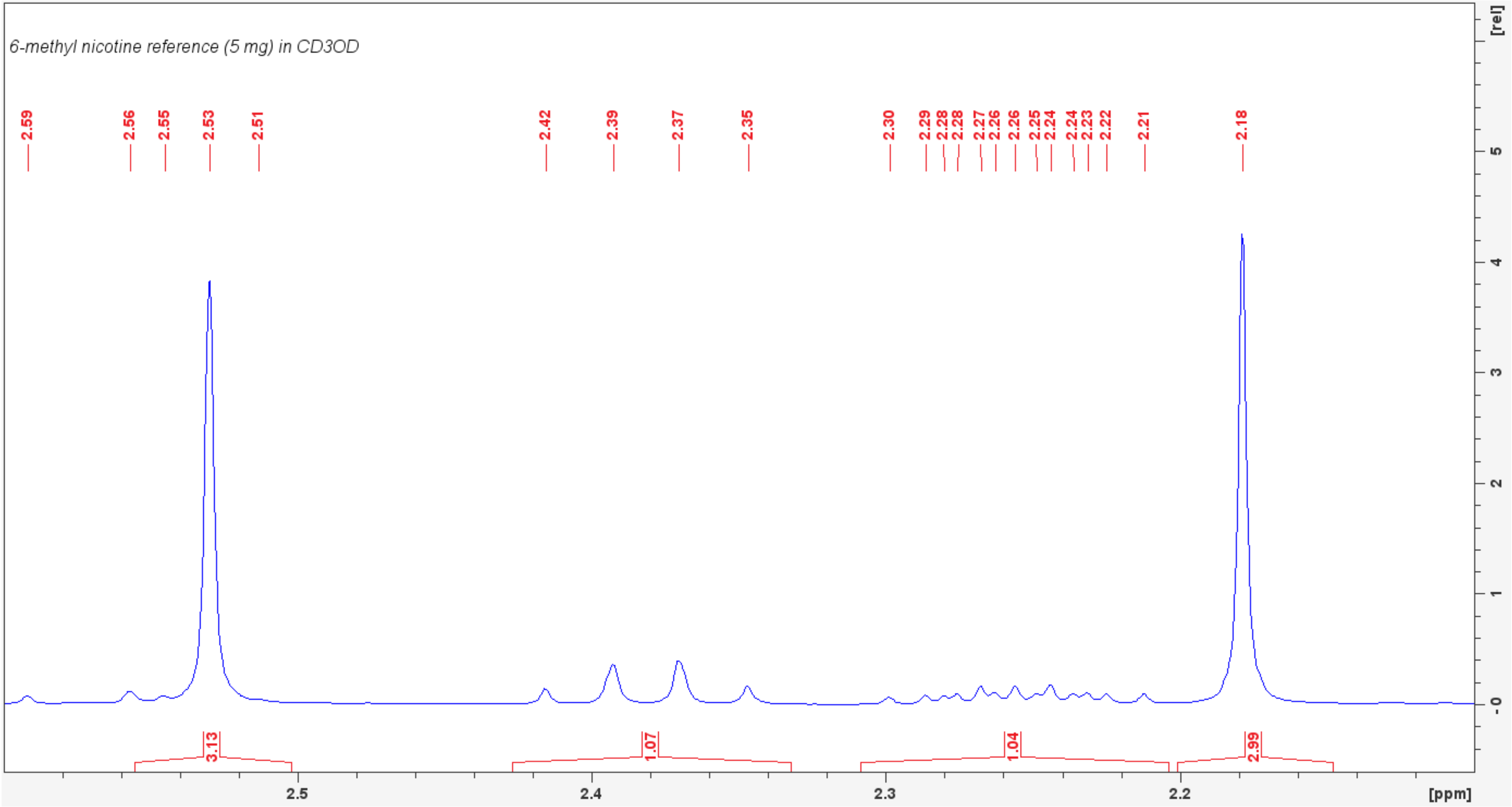
6-methyl nicotine reference (5 mg) in CD3OD
2.59
2.56
2.55
2.53
2.51
2.42
2.39
2.37
2.35
2.30
2.29
2.28
2.28
2.27
2.26
2.26
2.25
2.24
2.24
2.23
2.22
2.21
2.18
3.13
1.07
1.04
2.99
2.5
2.4
2.3
2.2
[ppm]
[rel]
5
4
3
2
1
0

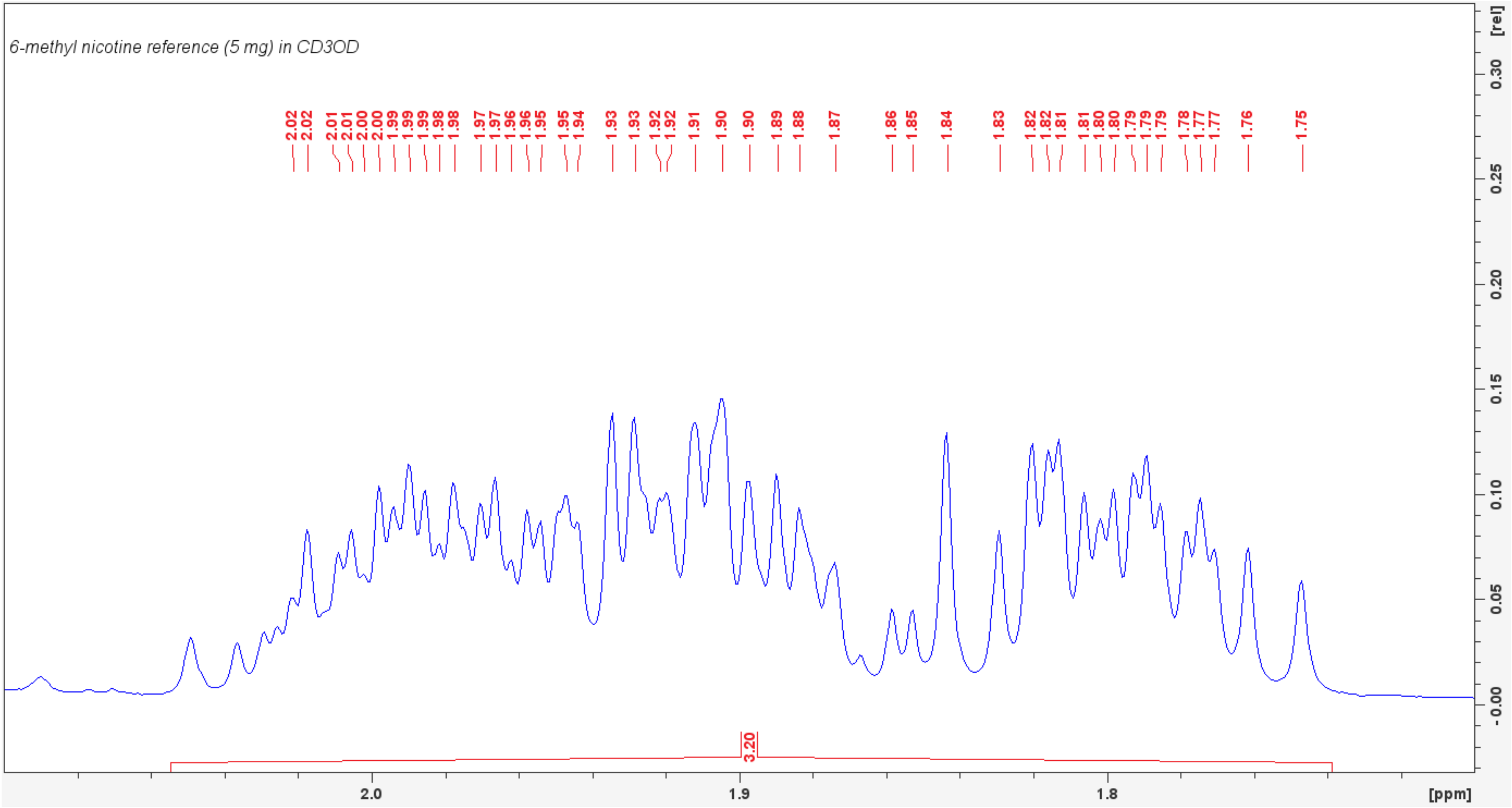

6-methyl nicotine reference (5 mg) in CD3OD
2.02
2.02
2.01
2.01
2.00
2.00
1.99
1.99
1.99
1.98
1.98
1.97
1.97
1.96
1.96
1.95
1.95
1.94
1.93
1.93
1.92
1.92
1.91
1.90
1.90
1.89
1.88
1.87
1.86
1.85
1.84
1.83
1.82
1.82
1.81
1.81
1.80
1.80
1.79
1.79
1.79
1.78
1.77
1.77
1.76
1.75
3.20
2.0
1.9
1.8
[ppm]
[rel]
0.30
0.25
0.20
0.15
0.10
0.05
-0.00

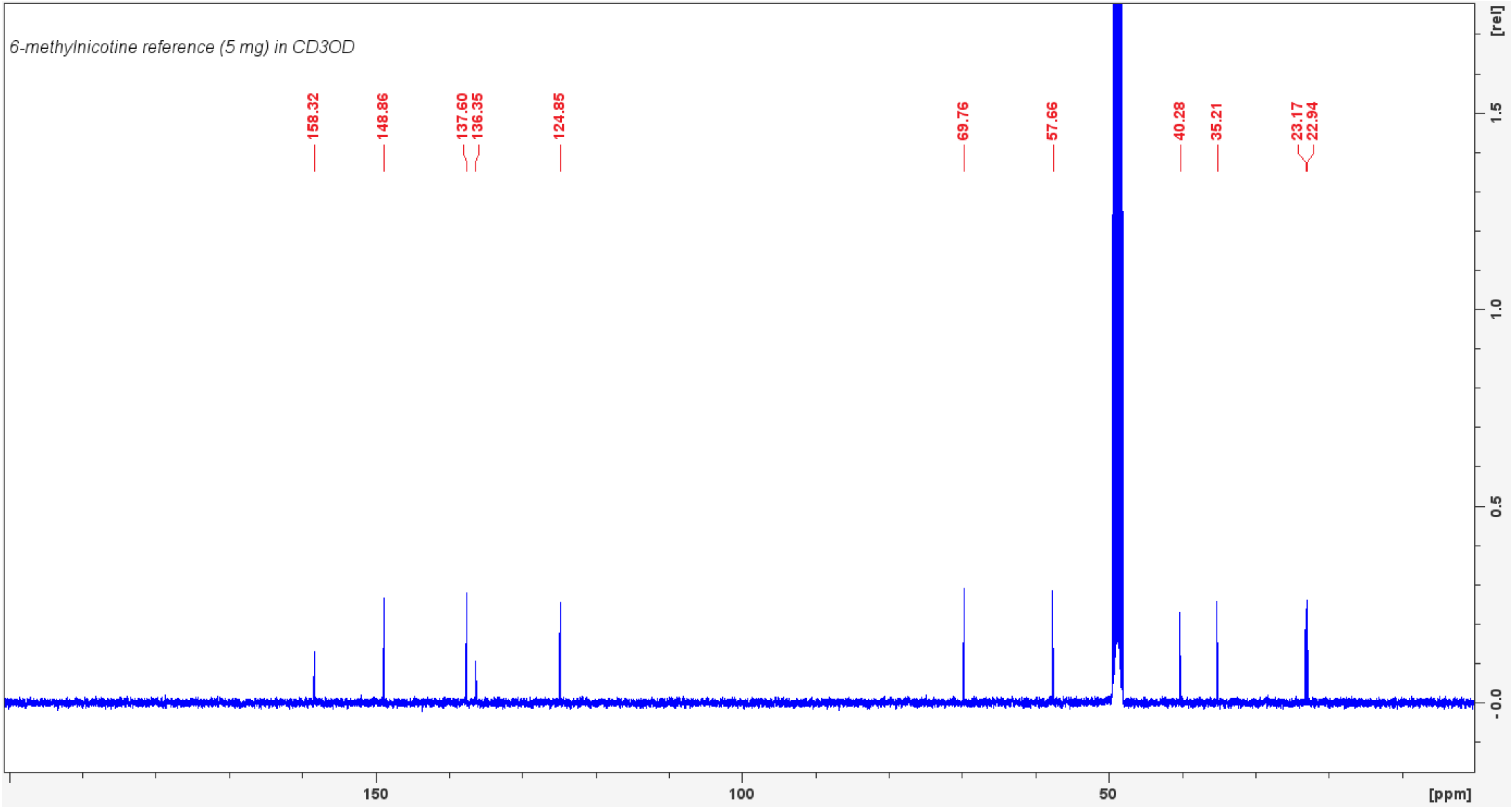

6-methylnicotine reference (5 mg) in CD3OD
158.32
148.86
137.60
136.35
124.85
69.76
57.66
40.28
35.21
23.17
22.94
[rel]
1.5
1.0
0.5
-0.0
150
100
50
[ppm]

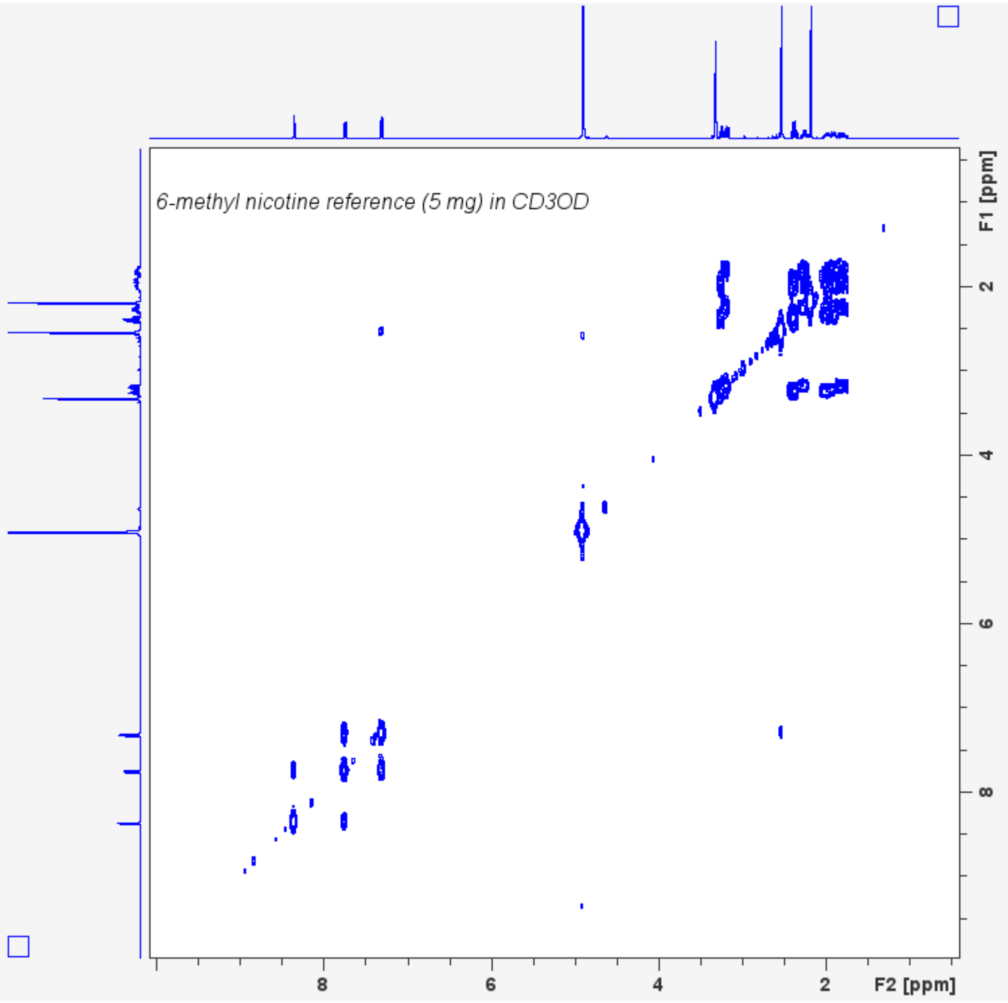

6-methyl nicotine reference (5 mg) in CD3OD
F1 [ppm]
2
4
6
8
8
6
4
2
F2 [ppm]

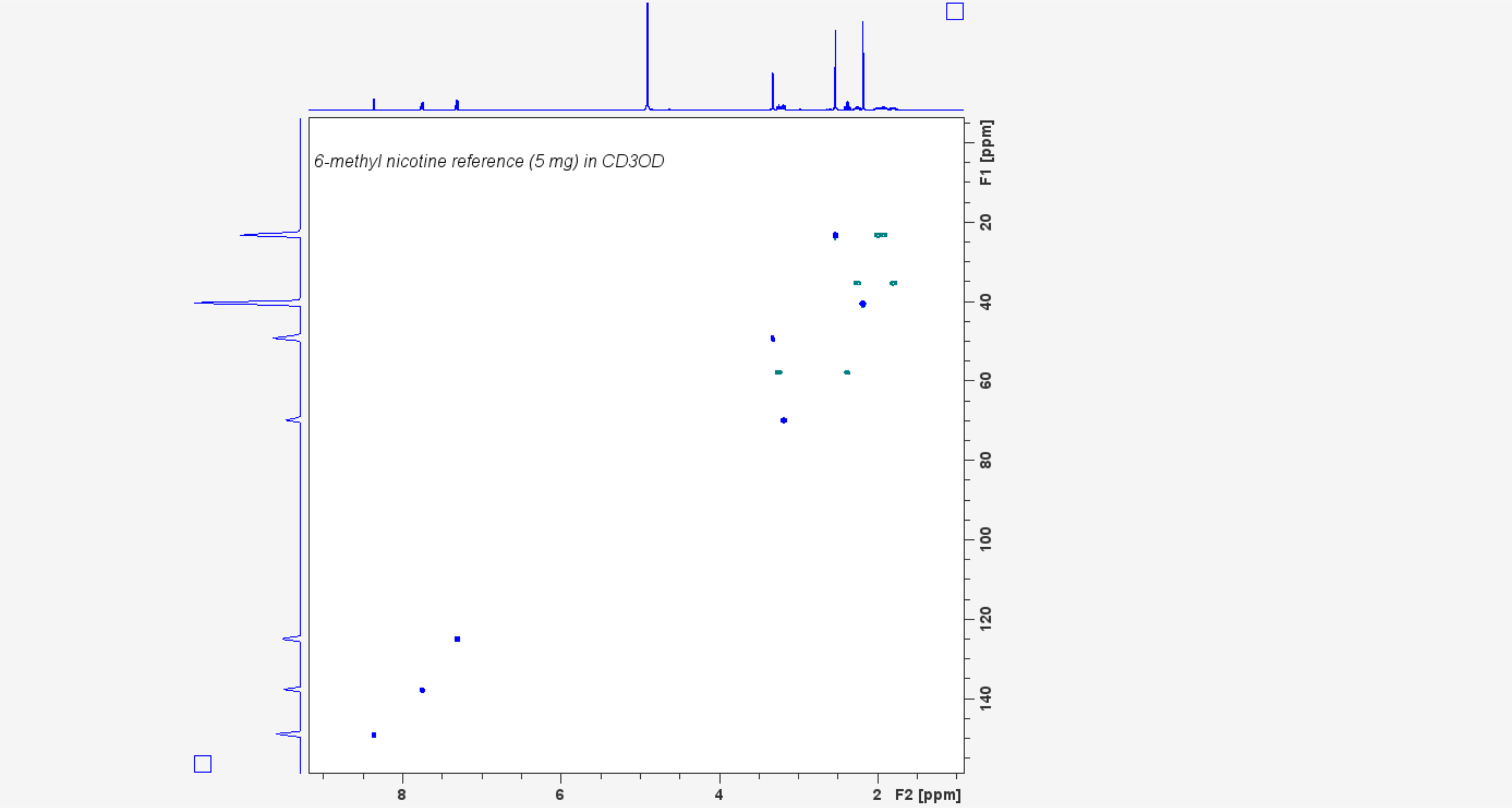

6-methyl nicotine reference (5 mg) in CD3OD
F1 [ppm]
20
40
60
80
100
120
140
8
6
4
2
F2 [ppm]

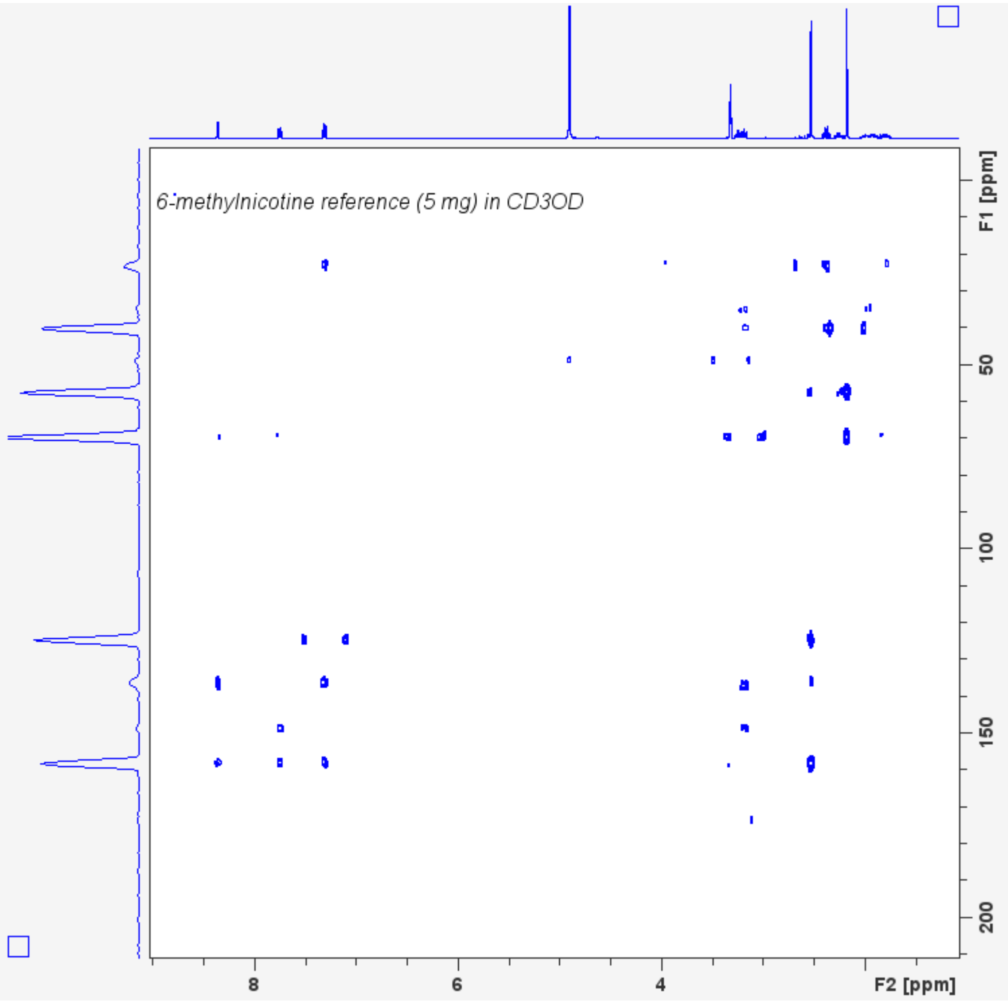

6-methylnicotine reference (5 mg) in CD3OD
F1 [ppm]
50
100
150
200
8
6
4
F2 [ppm]
