## Supplementary figures and images for "The emergence of a novel synthetic nicotine analog 6-methyl nicotine (6-MN) in proclaimed tobacco- and nicotine-free pouches available in Europe"

### Supplemental figure S20

10 µg/mL nicotine and 6-MN mixture

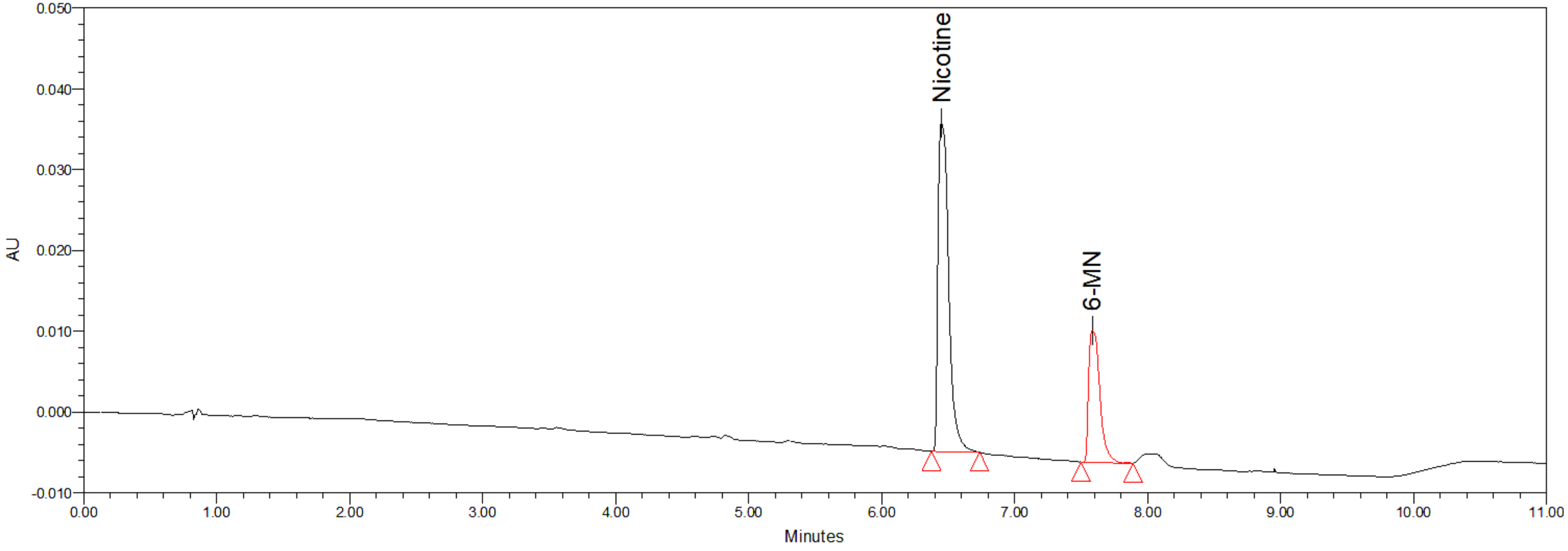


extract from pouches

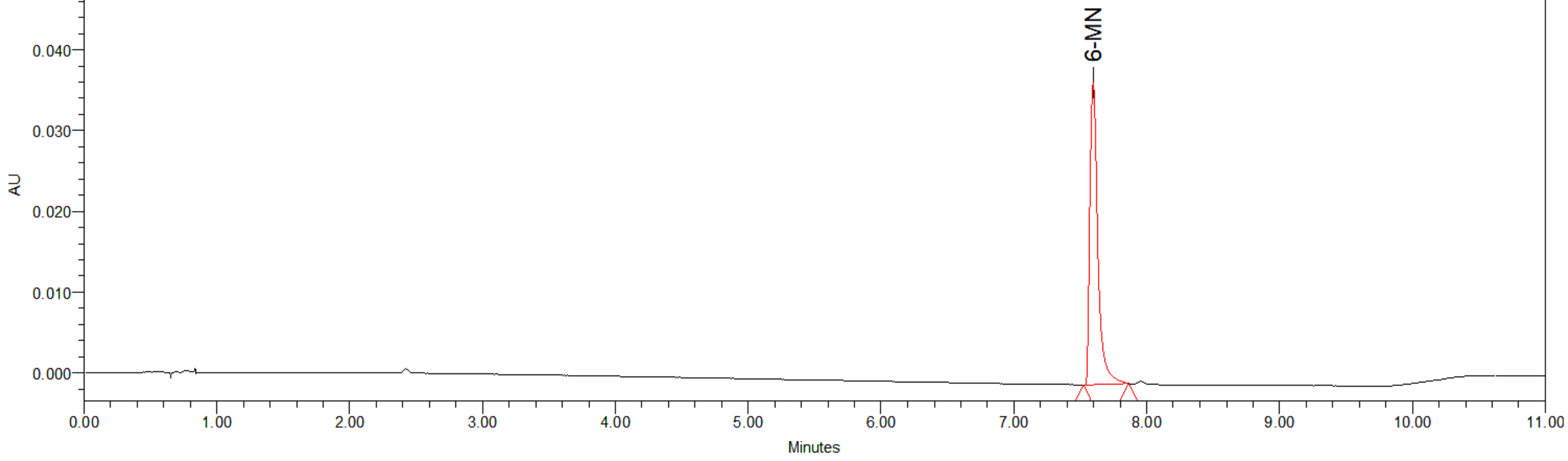
